# CoNVict: An Agentic AI System for Copy Number Variation Prioritization in Rare Disease Diagnosis

**DOI:** 10.64898/2026.03.16.26348493

**Authors:** Mert Gençtürk, Muti Kara, Furkan Özden

**Author notes:** These authors contributed equally to this work (Mert Gençtürk, Muti Kara).

## Abstract

Copy number variants (CNVs) are established contributors to rare genetic disorders, yet their clinical interpretation remains challenging in diagnostic genomics. Large CNVs frequently encompass multiple functional regions whose clinical significance can only be resolved in the context of the patient’s phenotype. Effective prioritization demands variant-level scoring of dosage sensitivity, structural consequences, and disease associations, and systematic comparison of candidates within the same clinical context. Current computational tools only partially address these requirements: they automate variant-level scoring but leave phenotype-guided evidence integration and cross-variant ranking to the clinician, creating a gap between annotation throughput and diagnostic decision-making. Agentic AI systems coordinate large language model-driven reasoning across structured multi-step pipelines and have shown strong performance on biomedical tasks requiring iterative evidence evaluation and contextual judgement, making them well suited to patient-specific variant interpretation where rigid scoring functions fall short. Here, we present `CoNVict`, a two-stage agentic AI system for patient-specific CNV prioritization. The system ranks CNVs through verdict classification that triages candidates and tournament ranking that performs pairwise comparisons via structured, in-context reasoning. Evaluated on simulated diagnostic cases spanning multiple clinical subspecialties, `CoNVict` substantially outperforms existing computational methods in identifying the causal CNV and maintains robust performance on variants of uncertain significance and non-coding variants without retraining. Our results demonstrate that agentic AI can bridge the gap between automated variant-level annotation and the patient-specific clinical reasoning required for CNV-driven genetic diagnosis. Availability and Implementation: Source code and data are available at https://github.com/Muti-Kara/CoNVict.

## Introduction

Rare diseases affect approximately 300 million people worldwide (Nguengang Wakap et al., 2020), and copy number variants (CNVs), deletions or duplications of 50 bp to megabases (Alkan et al., 2011), are a major genetic cause, accounting for 15–20% of cases with neurodevelopmental disorders or multiple congenital anomalies (Miller et al., 2010). However, unlike single nucleotide variations, CNVs present additional interpretive challenges. Spanning multiple genes, disrupting regulatory elements, unmasking recessive alleles, or creating gene fusions at breakpoints, their clinical impact depends on the dosage sensitivity of overlapping genes, the integrity of nearby regulatory architecture, and the patient’s phenotypic presentation (Rice and McLysaght, 2017; Spielmann et al., 2018). This makes CNV interpretation inherently a reasoning task that resists reduction to variant-level scoring and that early inter-laboratory studies showed to be highly inconsistent even among experts (Tsuchiya et al., 2009). The ACMG framework formalized this process through a quantitative scoring by pathogenicity (Riggs et al., 2020). Some tools, such as ClassifyCNV (Gurbich and Ilinsky, 2020) and AnnotSV (Geoffroy et al., 2018, 2021), automate its variant-level criteria while patient-specific criteria remain largely manual. Phenotype-agnostic methods like StrVCTVRE (Sharo et al., 2022), CADD-SV (Kleinert and Kircher, 2022), X-CNV (Zhang et al., 2021), and CNVoyant (Schuetz et al., 2024) can predict pathogenicity but cannot distinguish a pathogenic yet unrelated CNVs to the patient’s condition. However, phenotype-aware methods such as Exomiser (Smedley et al., 2015), PhenoSV (Xu et al., 2023) and SvAnna (Danis et al., 2022) integrate clinical, still these tools evaluates each CNV in isolation, therefore they can not perform the comparative evidence synthesis central to expert diagnostic reasoning.

Of reported CNVs in diagnostic sequencing, 10–15% are classified as variants of uncertain significance (VUS) (Nowakowska, 2017; Ravel et al., 2023). Periodic reanalysis can reclassify only few to actionable categories, leaving most unresolved. Novel and non-coding CNVs present particular challenges, as their functional consequences cannot be reliably inferred from existing annotations alone (Spielmann et al., 2018). These are precisely the cases where clinical reasoning, not pattern matching, determines diagnostic outcome.

Recent work on agentic AI systems demonstrates that multi-step, LLM-driven reasoning can meet this demand. DeepRare (Zhao et al., 2026) introduced a multi-agent system for rare disease differential diagnosis with transparent, evidence-linked reasoning, and the Deep Agentic Variant Prioritisation framework (Kara et al., 2026) showed that a tournament-based architecture attains substantially improved performance on VUS and novel cases by emulating the iterative evidence gathering of clinical geneticists. These results confirm that CNV interpretation is naturally suited to agentic approaches.

Here, we present `CoNVict`, an agentic AI system for patient-specific CNV prioritization. `CoNVict` ranks candidate CNVs through two stages: CNVerdict classification that triages candidates and tournament ranking that performs pairwise comparisons using gene keywords, dosage sensitivity metrics, and structural impact data. We evaluate `CoNVict` on clinical benchmark data spanning multiple diagnostic sources and variant types, and demonstrate that it achieves substantial improvements in recall over existing methods while generalising across clinical subspecialities and maintaining robust performance on VUS and non-coding variants through agentic reasoning.

## Results

### Overview of CoNVict

The `CoNVict` system is a multi-stage pipeline progressively filtering and ranking CNVs with LLM-based analyses. Starting from a VCF file, variants are first annotated by AnnotSV (Geoffroy et al., 2018). CNVoyant (Schuetz et al., 2024) then generates a preliminary ranking and selects a subset of candidates, the top 64 for WES simulation and top 512 for WGS. These candidate subsets are then processed by the CNVerdict stage, which utilizes an LLM to perform a three-class classification, “relevant,” “abstain,” or “irrelevant”, based on association with the patient’s phenotypes. Following this classification, the remaining variants, the top 32 for WES simulation and top 64 for WGS simulation, enter final tournament, where the LLM conducts pairwise comparisons that integrate detailed variant reports with the clinical context to generate a final, ranked list of the variants, top 4 for WES simulation and top 8 for WGS simulation.

### Prioritization Generalizes Across Diagnostic Sources and Sequencing Modalities

We evaluated `CoNVict` against Exomiser, PhenoSV, and SvAnna in 397 diagnostic cases with known causal CNVs compiled from ClinVar (Landrum et al., 2014), DECIPHER (Bragin et al., 2014), the Undiagnosed Diseases Network (Alsentzer et al., 2023; Ramoni et al., 2017), and novel, computationally derived cases.

`CoNVict` ranked the causal CNV at position 1 in 74.3% of cases and within the top 5 in 89.2% (Figure 2A), representing improvements of 10.3 and 7.3 percentage points over the next best method (Exomiser) at top-1 and top-5, respectively. At top-10, `CoNVict` reaches 92.9% compared to 88.9% for Exomiser. PhenoSV achieves moderate recall (37.5% top-1), while SvAnna remains below 18% at top-5.

**Figure 1.**
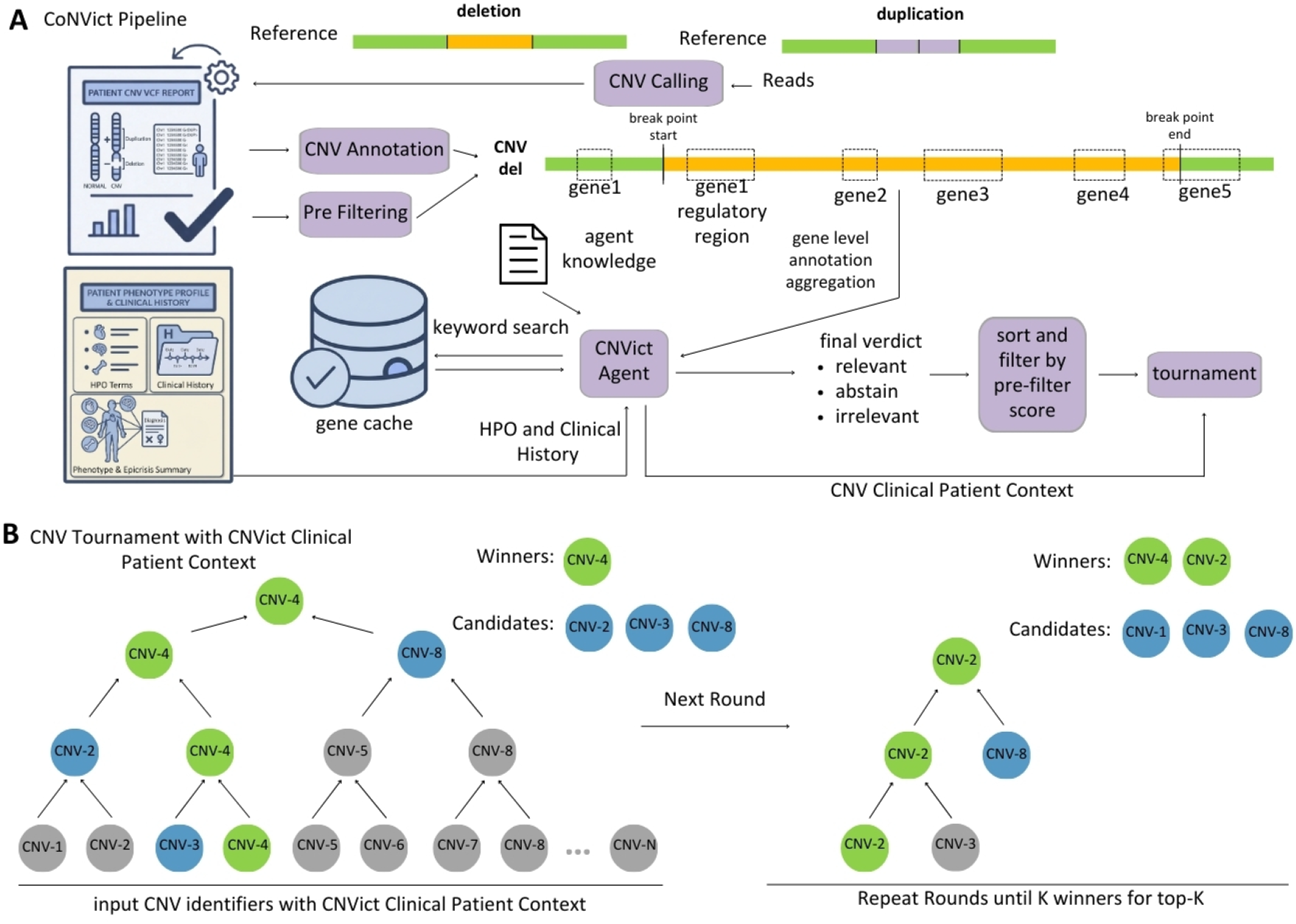
`CoNVict` **system overview. (A)** Core `CoNVict` pipeline integrating multiple components, such as AnnotSV to obtain gene-level annotations, a pre-filtering step retains the top-scoring candidates by CNVoyant score, a precomputed gene cache consists of phenotype-relevant keywords retrieved from literature, the `CoNVict` Agent aggregating annotations alongside the patient’s HPO terms and epicrisis to form the CNV Clinical Patient Context, CNVerdict assigning each variant a verdict of *relevant, abstain*, or *irrelevant* based on phenotypic fit, and last, the final tournament. **(B)** The tournament ranks candidates through iterative rounds of pairwise LLM comparisons conditioned on the `CoNVict` Clinical Patient Context. In the first round, all input CNV identifiers are randomly paired and compared; CNV-4 emerges as the winner. A second tournament is then run over the candidates that lost exclusively to CNV-4, identifying CNV-2 as rank 2. If there are odd numbers of candidates, then one of them automatically passes. This procedure repeats, each time collecting variants that lost only to already-ranked candidates, until *K* winners are identified for the final top-*K* list.

**Figure 2.**
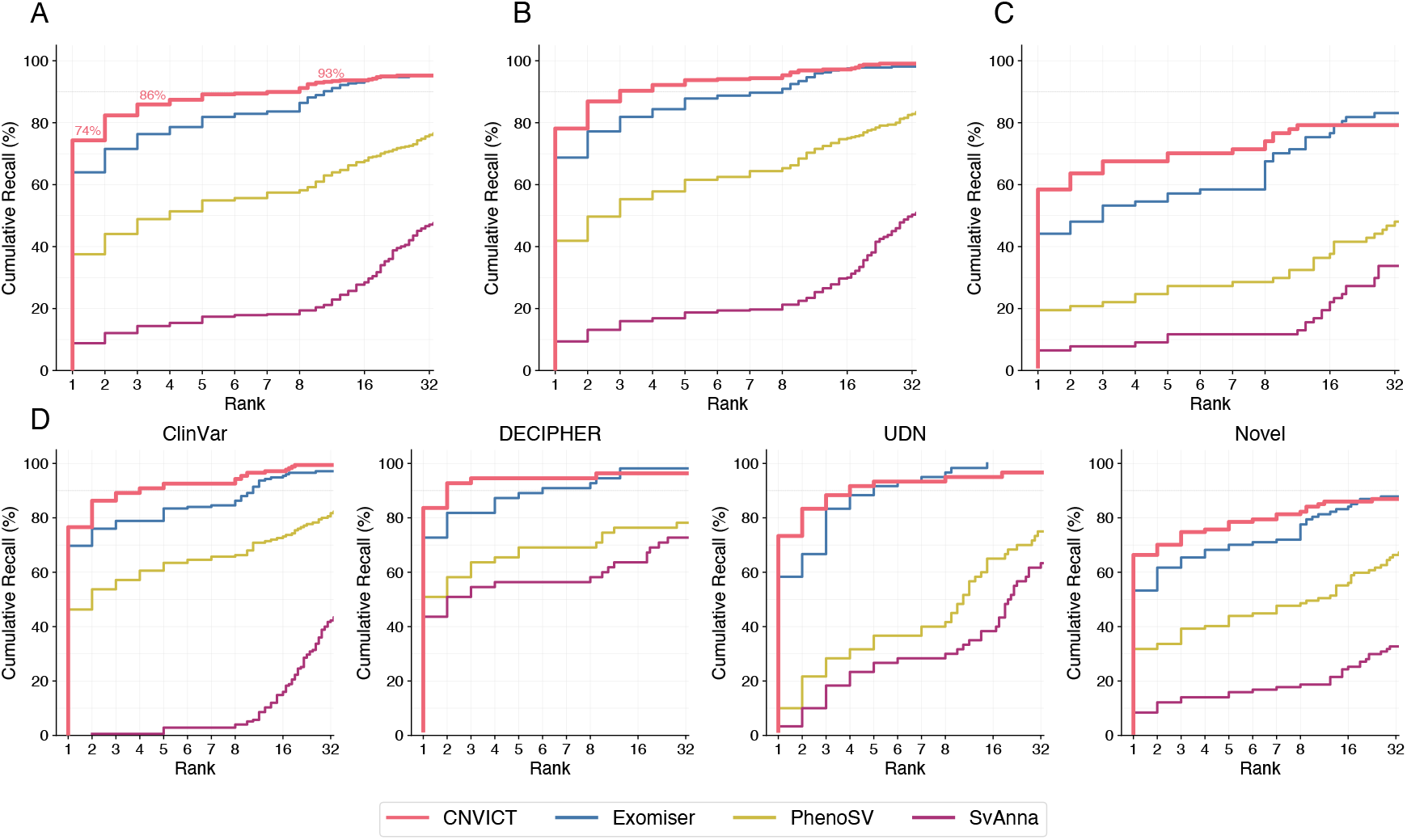
Cumulative distribution function (CDF) comparing diagnostic CNVs ranks for `CoNVict`, Exomiser, PhenoSV, and SvAnna across 397 simulated patients. The x-axis uses a piecewise-linear scale emphasizing clinically actionable ranks 1-8, with compressed intervals for ranks 9-16 and 17-32. **(A)** Overall performance across all simulated patients (n=397). **(B)** Performance on WES simulations (n=320) **(C)** Performance on WGS simulations (n=77) **(D)** Performance stratified by diagnostic CNV source: ClinVar (n=175), DECIPHER (n=55), UDN (n=60), Novel (n=107). Each CDF curve represents all evaluated samples within the respective subset.

When stratified by sequencing modality (Figure 2B,C), `CoNVict` achieves 78.1% top-1 recall in WES simulations (*n* = 320, mean 129 background CNVs per patient) and 58.4% in WGS simulations (*n* = 77, mean 969 background CNVs per patient), exceeding Exomiser by 9.4 and 14.3 percentage points, respectively. Despite the approximately 7.5-fold increase in background noise from WES to WGS, `CoNVict` maintains the highest recall across all rank thresholds. The performance gap between `CoNVict` and competing methods widens in the WGS setting, where PhenoSV top-5 recall drops from 61.6% to 27.3% and SvAnna from 18.8% to 11.7%, while `CoNVict` retains 70.1% top-5 recall.

Performance varies across data sources (Figure 2D). `CoNVict` achieves the highest top-1 recall on DECIPHER cases (83.6%, *n* = 55), followed by ClinVar (76.6%, *n* = 175) and UDN (73.3%, *n* = 60). In novel variants (n=107), absent from both ClinVar and DECIPHER (see Section 3.1), all methods show reduced recall, yet `CoNVict` still leads at 66.4% top-1 and 78.5% top-5. Detailed breakdowns by SV type (Supp. Fig. S5, Tab. S1), size category (Supp. Fig. S6, Tab. S2), and structural mechanism (Supp. Tab. S3) are provided in Supplementary Section 8. Against phenotype-agnostic scoring tools that rank variants without patient context, `CoNVict` also achieves the higher recalls, outperforming CNVoyant, AnnotSV, and StrVCTVRE across all rank thresholds (Supp. Fig. S15, Tab. S12). Further analysis of UDN cases, including the effect of distractor CNV burden on ranking accuracy, is provided in Supplementary Figures S13–S14.

### Agentic Reasoning Enables Prioritization Beyond Known Pathogenicity

We assessed `CoNVict` on two complementary definitions of variant uncertainty: database-level VUS annotations from ClinVar and UDN (*n* = 65), and variants classified as VUS by AnnotSV’s automated ACMG/ClinGen scoring (*n* = 124), which includes both literature-derived and computationally derived novel cases (Methods 3.1).

On database-annotated VUS cases (Figure 3A,B), `CoNVict` achieves 61.5% top-1 and 87.7% top-5 recall, compared to 58.5% and 78.5% for Exomiser, and wins the head-to-head comparison in 33.8% of cases versus 20.0% (Supp. Tab. S8 and S9). On the AnnotSV-classified VUS set (Figure 3C,D), `CoNVict` reaches 54.8% top-1 and 75.8% top-5 recall, exceeding Exomiser by 16.1 and 17.7 percentage points.

**Figure 3.**
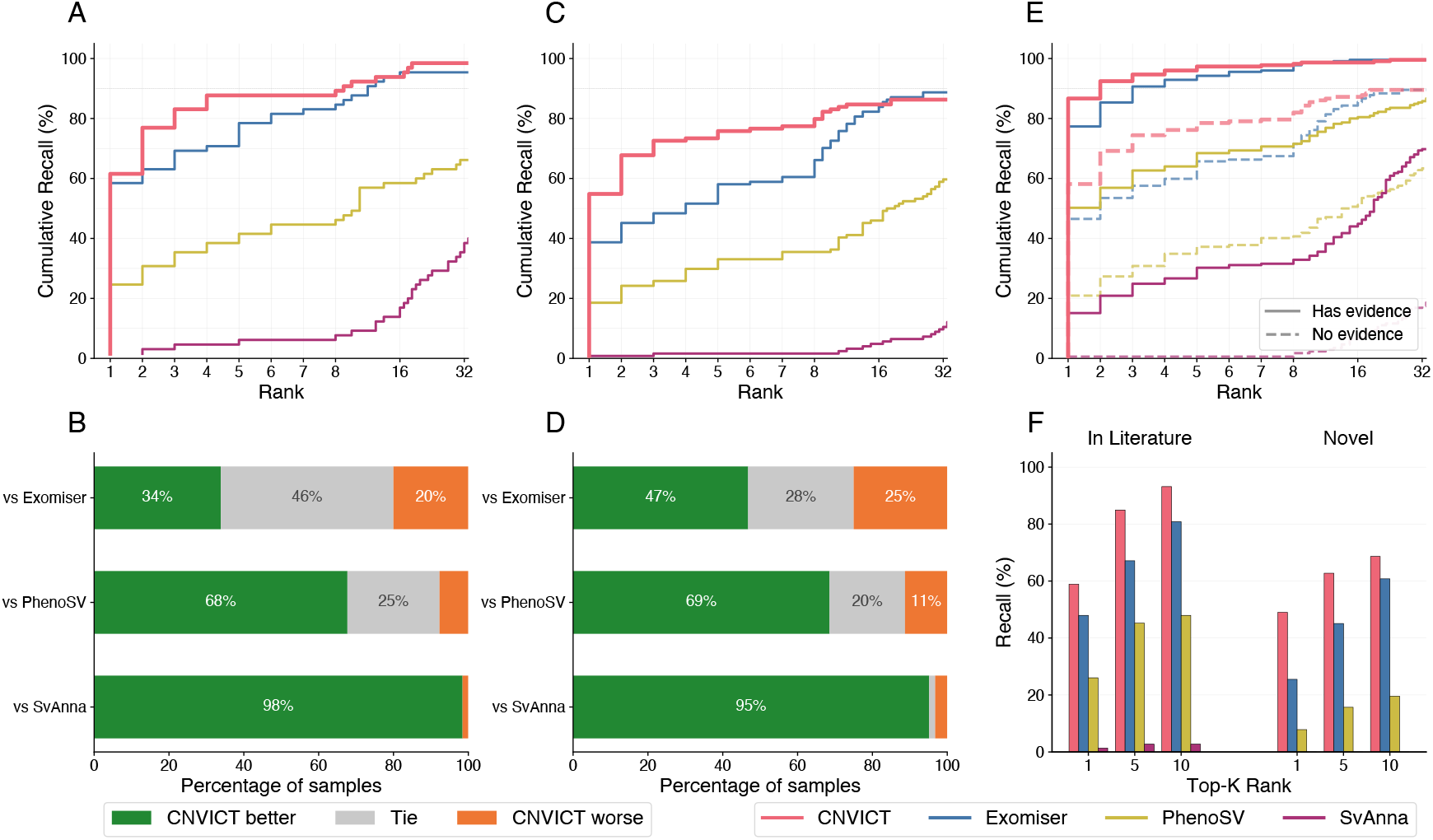
Prioritization performance on variants of uncertain significance. All panels compare `CoNVict`, Exomiser, PhenoSV, and SvAnna. **(A)** Diagnostic rank CDF for ClinVar and UDN VUS-classified variants (*n* = 65). **(B)** Head-to-head comparison of `CoNVict` versus each competitor on the VUS subset. **(C)** Diagnostic rank CDF for variants classified as VUS by AnnotSV (*n* = 124). **(D)** Head-to-head comparison for the AnnotSV-classified VUS subset. **(E)** Diagnostic rank CDF for all 397 cases stratified by presence of prior pathogenic SV evidence (solid = has evidence, *n* = 225; dashed = no evidence, *n* = 172; see text for definitions). **(F)** Top-*K* recall at ranks 1, 5, and 10 for AnnotSV-classified VUS variants, separated by origin: literature-derived (*n* = 73) versus novel (*n* = 51).

To assess how prior knowledge influences prioritization, we stratified all 397 cases by overlaps with any reported pathogenic SV or established disease-associated gene (Figure 3E), using AnnotSV’s P loss source and P gain source fields (Geoffroy et al., 2018). These fields aggregate evidence from ClinVar (Landrum et al., 2014), dbVar (Lappalainen et al., 2013), OMIM morbid genes (Amberger et al., 2019), and ClinGen dosage sensitivity curations (Rehm et al., 2015). Variants with at least one prior pathogenic record were assigned to the “has evidence” stratum (*n* = 225); the remainder (*n* = 172) have none. All methods show reduced performance without prior evidence, but `CoNVict` retains the highest absolute recall in both strata, with a top-1 drop of 28.5 percentage points (86.7% to 58.1%) compared to 30.8 for Exomiser and 29.3 for PhenoSV. Performance stratified by ClinVar review star rating is shown in Supplementary Figures S11 and S12.

Finally, among AnnotSV-classified VUS variants separated by origin (Figure 3F), `CoNVict` achieves 49.0% top-1 and 62.7% top-5 recall on novel cases (*n* = 51), compared to 25.5% and 45.1% for Exomiser. PhenoSV drops to 7.8% top-1 and SvAnna fails to rank any novel variant within the top 1. Detailed recall stratified by literature evidence status and pairwise tool comparisons on the novel subset are reported in Supplementary Tables S11 and S10. `CoNVict` maintains the highest recall across all diagnostic source subcategories (Supp. Fig. S8, Table S5), disease systems (Supp. Fig. S9, Table S6), and ClinGen dosage sensitivity tiers (Supp. Fig. S10, Table S7).

### In-Context Reasoning Maintains Prioritization Across Variant Localization

To assess how variant localization affects prioritization performance, we stratified the benchmark set into coding variants (*n* = 334), defined as CNVs overlapping at least one exon of a protein-coding gene, and non-coding variants (*n* = 63), comprising intronic and intergenic CNVs with no exonic overlap. Figure 4A shows cumulative recall as a function of rank for coding variants, where all tools achieve their highest performance. Figure 4B presents the corresponding curves for non-coding variants, where all four tools exhibit markedly lower recall. `CoNVict`’s top-1 recall decreases from 79.9% on coding variants to 44.4% on non-coding variants, a reduction of 35.5 percentage points. The remaining tools show steeper declines: Exomiser drops from 74.0% to 11.1%, PhenoSV from 44.3% to 1.6%, and SvAnna from 10.5% to 0.0%.

**Figure 4.**
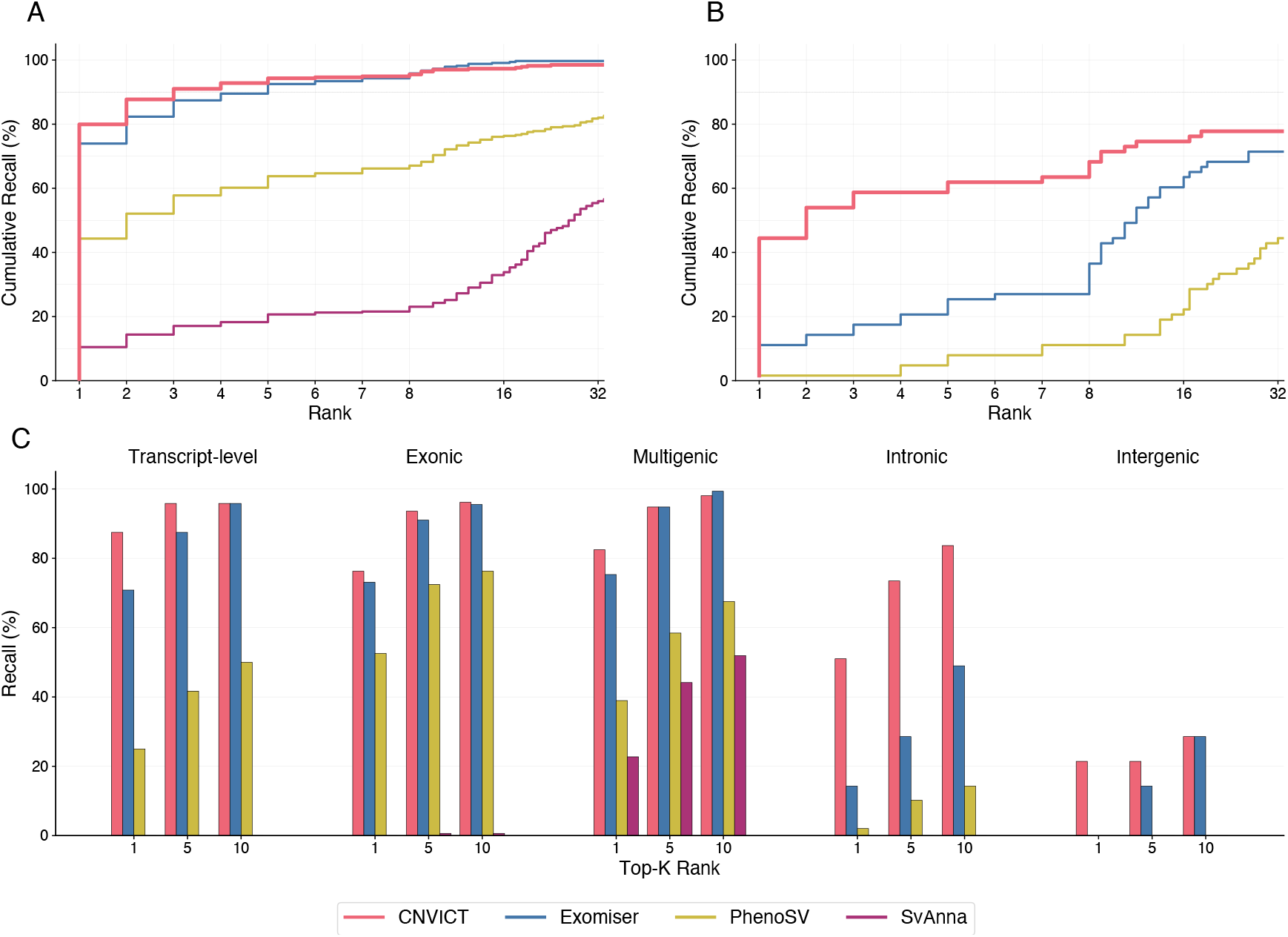
**(A)** CDF plot for coding variants (n = 334). **(B)** CDF plot for non-coding variants (n = 63). **(C)** Top-1, top-5, and top-10 recall stratified by SO consequence category. Transcript-level (n = 24): whole-gene deletions or duplications. Exonic (n = 156): Partial exon loss or gain. Multigenic (n = 154): CNVs spanning multiple genes. Intronic (n = 49): CNVs located entirely within intronic sequence. Intergenic (n = 14): CNVs in intergenic or regulatory regions with no direct gene overlap.

Figure 4C further stratifies variants by Sequence Ontology (SO) (Eilbeck et al., 2005) consequence. Coding categories (transcript-level, multigenic, exonic) yield the highest recall, with `CoNVict` achieving 87.5%, 82.5%, and 76.3% top-1 respectively. Performance diverges for non-coding variants: for intronic variants (n=49), which reside within gene boundaries but do not disrupt exonic sequence, `CoNVict` attains 51.0% top-1 recall, compared to 14.3% for Exomiser and 2.0% for PhenoSV, while SvAnna fails to rank any intronic variant first. On intergenic variants (*n* = 14), `CoNVict` is the only tool with any top-1 recall (21.4%). Full recall across all rank thresholds for each SO category is reported in Supplementary Table S4.

## Methodology

### Simulation Benchmark Construction

We constructed a benchmark of 397 simulated diagnostic cases, each comprising a single causal CNV spiked into a realistic background of population-level structural variants. Cases were split into 320 WES and 77 WGS simulations. A separate 38-case development set, constructed identically, was reserved for prompt tuning and excluded from all evaluations.

Background CNV profiles were drawn from the 1000 Genomes Project Phase 3 structural variant call set (The 1000 Genomes Project Consortium, 2015; Sudmant et al., 2015), which combined multiple discovery algorithms across 2,504 individuals from 26 populations. Each patient was assigned a unique 1000G sample. WES backgrounds retained only exon-overlapping CNVs, while WGS backgrounds retained the full structural variant profile. Any background variant overlapping the spiked-in causal CNV was removed to prevent confounding.

Causal CNVs were drawn from six categories spanning a range of clinical difficulty: pathogenic, likely pathogenic, and VUS entries from ClinVar (Landrum et al., 2014), each requiring gene annotation and a mapped OMIM disease (Amberger et al., 2019); curated CNV syndrome regions from DECIPHER (Bragin et al., 2014); gene–disease associations validated by the Undiagnosed Diseases Network (UDN) (Alsentzer et al., 2023; Ramoni et al., 2017); and computationally derived CNVs placed over ClinGen dosage-sensitive genes (Rehm et al., 2015). Computationally derived CNV’s include both coding variants over genes lacking any existing structural variant in ClinVar and non-coding variants in intronic or upstream regulatory regions verified against GENCODE v44 (Frankish et al., 2021). UDN cases were split equally between genes with no prior gene–disease association in ClinVar, ClinGen, or OMIM (novel associations) and genes already catalogued in at least one of these resources (known associations), and additionally included distractor gene CNVs in the background as realistic diagnostic noise. ClinVar and DECIPHER entries underwent randomized breakpoint perturbation to prevent trivial identification via exact coordinate matching against source databases. For each non-UDN case, a synthetic clinical narrative was generated by querying the causal gene–disease pair via web retrieval, synthesizing a patient epicrisis with an LLM, and extracting up to five standardized HPO terms (Köhler et al., 2021) through retrieval-augmented generation. UDN patients retained their original clinician-validated terms.

Full details of data sources, CNV generation procedures, perturbation strategies, background SV construction, and the phenotype extraction pipeline are provided in Supplementary Section 13.

## Method

### Problem Definition

Let *V* be the set of observed CNVs, *P* the patient’s HPO terms, and *c* the clinical notes. Each CNV *v*_*i*_ ∈ *V* has a feature vector **x**_*i*_ containing genomic coordinates, type, length, overlapping genes, gene-level pathogenicity scores, associated phenotypes and diseases, population frequencies, and ACMG classification. Let *G* be the set of genes covered by *V*, and let **X** denote the stacked feature matrix. The input is (*V, P, c*, **X**, *G*). The goal is to identify a causative subset *V* ^*⋆*^ ⊆ *V* and output a ranked list *v*(1) ≻ *v*_(2)_ ≻ … ≻ *v*_(|*V ⋆*|)_.

### Filtering

To reduce the cost of downstream LLM inference, we rank all candidate CNVs using CNVoyant (Schuetz et al., 2024) pathogenicity scores and retain the top *N*_WES_ = 64 for WES and *N*_WGS_ = 512 for WGS samples. These cutoffs are not arbitrary: These cutoffs retain ¿96% of causal CNVs (Supp. Tab. X) while halving the candidate set. More in-depth analysis of strong diminishing returns exists in Supplementary Figure S19) and Supplementary Table S18 alongside larger *k* yields marginal recall gains at substantially higher LLM cost. Notably, the pipeline is agnostic to the specific pre-filtering method: any score-based ranking that enriches pathogenic variants toward the top of the list, such as the AnnotSV or Exomiser scores, can serve as a drop-in replacement (Supp. Fig. S19). The resulting variant set *V* ^*′*^ proceeds to the Verdict stage.

### Gene Cache

To enable efficient gene-level reasoning, we precompute a gene cache for all genes obtained through large-scale web retrieval and language model grounding. Each gene report has information from OMIM (Amberger et al., 2019), HPO (Köhler et al., 2021), ClinVar (Landrum et al., 2014) (for full list look at Supplementary Figure S17)). Then, for each gene *g*, an LLM generates a compact set of phenotype- and disease-relevant keywords *k*(*g*) from this report, capturing its core clinical associations (Supp. Fig. S18). At inference time, given a variant *v*_*i*_ mapping to gene *g*, we retrieve *k*(*g*) as the gene context *c*(*v*_*i*_). This compacted cache has 14.63 keywords per gene on average, keeping context manageable for CNVs spanning 50+ genes, allowing LLMs to process multi-gene variants without overflowing context windows. Full details of web-retrieval and language grounding alongside statistics of compacted gene cache are provided in Supplementary Table S14, S15, S16 and Figure S17.

### CNVerdict

After initial filtering, we apply an LLM-based triage step, termed CNVerdict, to assess the clinical relevance of each remaining variant. For every *v*_*i*_ ∈ *V* ^*′*^, the system constructs a structured prompt (Supp. Fig. S21) from four elements: the variant’s AnnotSV annotation vector **x**_*i*_, phenotype keywords from the gene knowledge cache *c*(*g*) for each overlapping gene *g* ∈ *G*_*i*_, the patient’s HPO terms *P*, and the clinical epicrisis *e*. A single LLM call (Supp. Tab. S19) classifies the variant into *y*_*i*_ ∈ (relevant, abstain, irrelevant) according to the evaluation criteria detailed in the prompt template (Supp. Fig. S21). Because each variant is assessed independently, all |*V* ^*′*^| calls are issued in a single parallel batch (Supp. Tab. S21). Variants are then re-ranked with priority order relevant ≻ abstain ≻ irrelevant; within each class, the original CNVoyant score 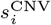 serves as a secondary sort key. From this re-ordered list *V* ^*′′*^, we retain the top *M*_WES_ = 32 for WES and *M*_WGS_ = 64 for WGS, yielding the reduced set *V* ^*′′′*^ that proceeds to the tournament stage.

### Detailed Variant Reports

For tournament variants, we expand gene context to include regulatory targets. Using regulatory annotations, we identify 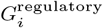 . The complete gene set is 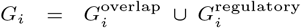. The detailed report 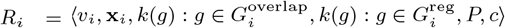 explicitly separates the two groups, providing the LLM with richer context about both direct coding disruption and potential regulatory effects during pairwise comparisons.

### Tournament

The final prioritization step applies tournament-based sorting to the reduced variant set *V* ^*′′′*^ (top 32 for WES, top 64 for WGS). Pairs are formed by randomly matching variants, and for each pair (*v*_*i*_, *v*_*j*_ ) the LLM compares them conditioned on the patient’s HPO terms *P*, clinical epicrisis *e*, and the two CNV representations (Supp. Fig. S20) according to the evaluation criteria in the comparison prompt (Supp. Fig. S22). To leverage the existing ranking, the higher-ranked variant from CNVerdict is presented as CNV 1 in the prompt, exploiting the model’s positional consistency bias. Unpaired variants in odd-sized rounds advance automatically.

To extract the top-*K* variants (*K*_WES_ = 4, *K*_WGS_ = 8), we first find first winner which is rank 1. For each subsequent rank *r* ∈ (2, …, *K*), we run a sub-tournament over all variants that lost directly to any current top-(*r*−1) variants that has not ranked yet. This ensures each ranked variant is the strongest among those not yet eliminated by a higher-ranked candidate.

The total number of LLM calls and sequential steps across both stages is analyzed in Supplementary Table S20 and S21. The final top-*K* list is returned with comparison histories for interpretability, along with a clinician-facing summary aggregating the detailed reports of top candidates. The independent contribution of each stage is evaluated in the ablation study (Supp. Fig. S16, Table S13).

## Discussion

CNV interpretation presents a fundamentally different challenge from single nucleotide variant analysis. A single CNV can span dozens of genes, partially disrupt coding sequences, delete or duplicate regulatory elements, and alter topologically associating domain boundaries, thereby affecting genes that lie entirely outside the variant’s coordinates through position effects (Spielmann et al., 2018). Tools such as AnnotSV faithfully collect this multi-layered annotation, but existing methods, whether phenotype-agnostic scorers or phenotype-aware tools such as Exomiser, PhenoSV, and SvAnna, apply static scoring functions that cannot determine which elements of a variant’s annotation are relevant to a specific patient’s presentation, a step the ACMG/ClinGen framework explicitly requires but leaves to the clinician (Riggs et al., 2020).

`CoNVict` addresses this gap by aggregating variant-level annotation into structured gene reports and evaluating them against patient HPO terms and clinical history through LLM-based reasoning. Across 397 simulated diagnostic cases, `CoNVict` ranked the causal CNV first in 74.3% of cases and within the top five in 89.2%, improving over Exomiser by 10.3 and 7.3 percentage points respectively. The performance gap widened in the WGS setting, where a 7.5-fold increase in background variants constitutes the most challenging noise environment, suggesting that patient-specific contextual reasoning becomes proportionally more valuable as the search space grows.

VUS represent 10–15% of reported CNVs, and periodic reanalysis reclassifies only a small minority to actionable categories (Ravel et al., 2023). On variants classified as VUS by AnnotSV, including novel cases absent from curated databases, `CoNVict` reached 54.8% top-1 recall versus 38.7% for Exomiser, with the margin widening further on novel VUS cases (49.0% vs. 25.5%). Current tools are predominantly gene-centric and provide limited means of interpreting regulatory impact (Danis et al., 2022), and on non-coding variants `CoNVict` achieved 44.4% top-1 recall compared to 11.1% for Exomiser and 0.0% for SvAnna, and was the only tool to rank any intergenic variant first. Both results reflect the ability of in-context reasoning to aggregate heterogeneous annotation and weigh it against phenotype, though absolute performance on these hardest cases remains well below well-annotated variants, underscoring how much interpretive difficulty persists across the field.

### Limitations and Future Directions

The benchmark relies on simulated cases with synthetic phenotype profiles rather than prospectively collected clinical patients, and performance on real diagnostic cohorts may differ. Background CNVs were drawn from population-level structural variant calls, which does not fully replicate the complexity of clinical genomes, including compound heterozygosity or co-occurring pathogenic variants. `CoNVict` does not incorporate inheritance or trio information, which clinical guidelines treat as informative evidence for variant classification (Riggs et al., 2020), and is not designed for on-target or pathogenicity scoring in isolation. LLM-based reasoning introduces computational costs that may limit throughput at clinical scale, and outputs inherit general LLM limitations including sensitivity to prompt formulation and potential run-to-run inconsistency. Non-coding performance, while improved relative to existing tools, remains constrained by the incompletely characterised state of regulatory annotation, a limitation shared across the field. Future work should evaluate `CoNVict` on prospective clinical cohorts, incorporate trio-based inheritance reasoning, and explore integration with functional genomic evidence to further improve prioritization of non-coding and novel variants.

## Data Availability

The CoNVict source code is available at https://github.com/Muti-Kara/CoNVict. All datasets used in this study are publicly available from sources cited in the manuscript.

## Author Contributions

FO designed the study. MK and MG designed and implemented the models. MK and MG performed the experiments. All authors approved the manuscript.

## Rights Retention Statement

This work is supported by Lidya Genomics. For the purpose of Open Access, the author has applied a CC-BY-NC-ND 4.0 International license to any Author Accepted Manuscript (AAM) version arising from this submission.

## Supplementary Materials: CONVICT

### Structural Variant Stratification

**Table S1.** Overall Top-*K* recall and stratification by structural variant (SV) type. Top-1, Top-5, Top-10, Top-16, Top-32, and Top-64 recall (%) for each of four prioritization tools (`CoNVict`, Exomiser, PhenoSV, SvAnna) across all 397 benchmark patients (“All”) and stratified by SV type: deletions (DEL, *n* = 321) and duplications (DUP, *n* = 76). Recall is defined as the percentage of patients for whom the diagnostic CNV was ranked at or above a given Top-*K* threshold. Accompanies Supplementary Figure S1.

| Stratum | Tool | Top-1 | Top-5 | Top-10 | Top-16 | Top-32 | Top-64 |
| --- | --- | --- | --- | --- | --- | --- | --- |
| All (*n* = 397) | CoNVict | 74.3 | 89.2 | 92.9 | 93.7 | 95.2 | 95.5 |
|  | Exomiser | 64.0 | 81.9 | 88.9 | 93.5 | 95.2 | 97.5 |
|  | PhenoSV | 37.5 | 54.9 | 61.0 | 67.8 | 76.1 | 88.2 |
|  | SvAnna | 8.8 | 17.4 | 20.4 | 28.5 | 47.1 | 62.5 |
| DEL (*n* = 321) | CoNVict | 76.0 | 90.0 | 93.5 | 94.4 | 96.3 | 96.6 |
|  | Exomiser | 65.1 | 81.6 | 88.8 | 93.8 | 95.3 | 98.1 |
|  | PhenoSV | 36.1 | 53.9 | 60.4 | 67.9 | 77.3 | 90.3 |
|  | SvAnna | 7.5 | 15.0 | 18.1 | 26.8 | 47.0 | 64.2 |
| DUP (*n* = 76) | CoNVict | 67.1 | 85.5 | 90.8 | 90.8 | 90.8 | 90.8 |
|  | Exomiser | 59.2 | 82.9 | 89.5 | 92.1 | 94.7 | 94.7 |
|  | PhenoSV | 43.4 | 59.2 | 63.2 | 67.1 | 71.1 | 78.9 |
|  | SvAnna | 14.5 | 27.6 | 30.3 | 35.5 | 47.4 | 55.3 |

**Figure S5.**
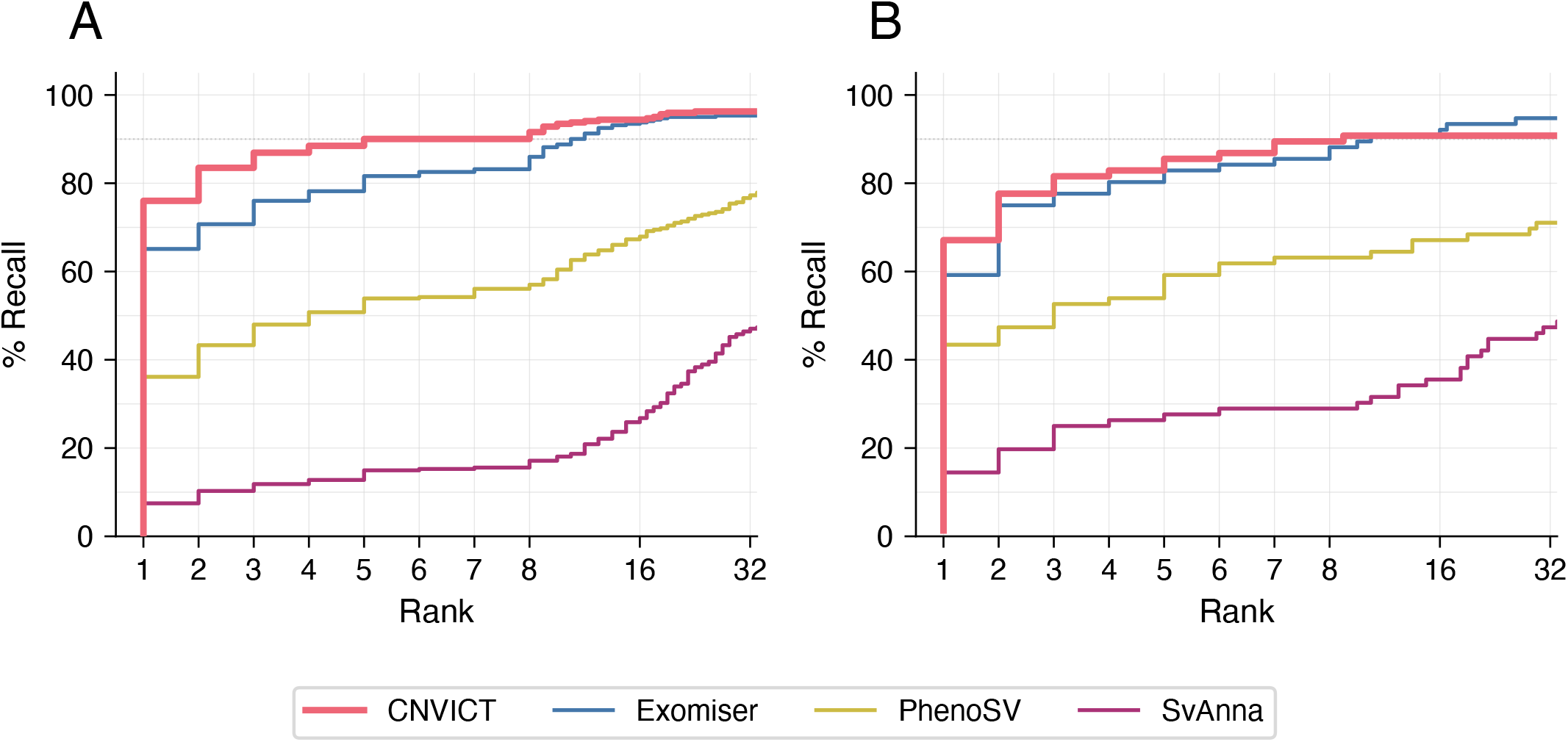
Cumulative distribution of diagnostic CNV ranking stratified by SV type. **(A)** Deletions. **(B)** Duplications. Each curve represents the cumulative percentage of simulated patients for whom the diagnostic CNV was ranked at or above a given rank threshold by each prioritization tool. Ranks are displayed on a piecewise-linear scale that is linear from 1 to 8 and logarithmically compressed beyond rank 8, placing visual emphasis on performance at the top of the ranked list. Four prioritization tools are compared: `CoNVict`, Exomiser, PhenoSV, and SvAnna. The grey dotted line marks 90% recall. A shared legend is provided at the bottom of the figure. See Supplementary Table S1 for Top-1/5/10/16/32/64 recall per tool and SV type.

**Figure S6.**
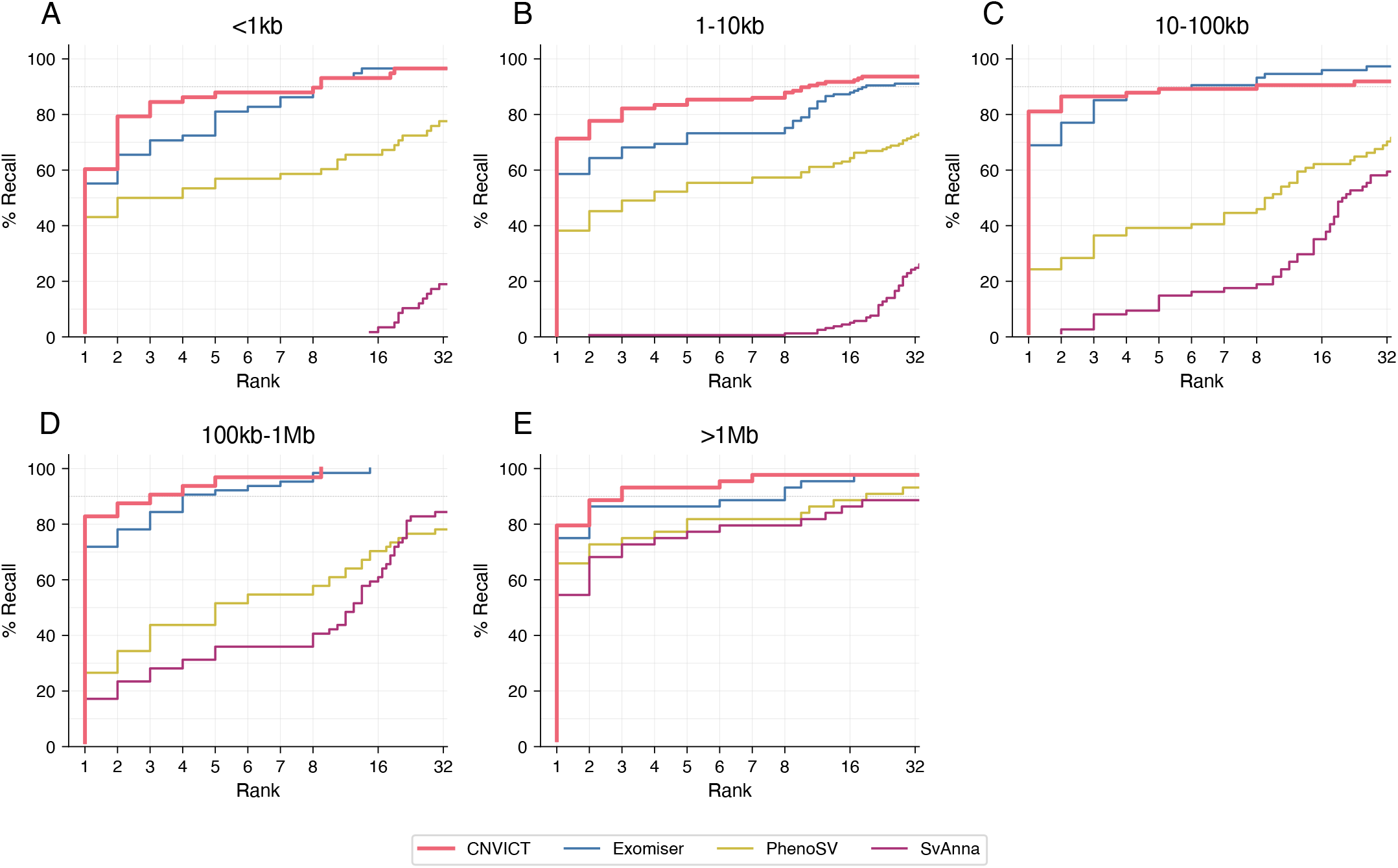
Cumulative distribution of diagnostic CNV ranking stratified by CNV size category. **(A)** *<*1 kb. **(B)** 1–10 kb. **(C)** 10–100 kb. **(D)** 100 kb–1 Mb. *>*1 Mb. Each panel shows the CDF of diagnostic CNV rank for all four tools within a given size category. The piecewise-linear rank scale compresses higher ranks to emphasize performance at the top of the ranked list. The grey dotted line marks 90% recall. Empty panels (if any size category contains no patients) are hidden. A shared legend is provided at the bottom of the figure. See Supplementary Table S2 for Top-1/5/10/16/32/64 recall per tool and size category.

**Table S2.** Top-*K* recall by CNV size category. Top-1, Top-5, Top-10, Top-16, Top-32, and Top-64 recall (%) for each of four prioritization tools (`CoNVict`, Exomiser, PhenoSV, SvAnna), stratified by five size categories: *<*1 kb (*n* = 58), 1–10 kb (*n* = 157), 10–100 kb (*n* = 74), 100 kb–1 Mb (*n* = 64), and *>*1 Mb (*n* = 44). Accompanies Supplementary Figure S2.

| Size | Tool | Top-1 | Top-5 | Top-10 | Top-16 | Top-32 | Top-64 |
| --- | --- | --- | --- | --- | --- | --- | --- |
| <1 kb (*n* = 58) | CoNVict | 60.3 | 87.9 | 93.1 | 93.1 | 96.6 | 96.6 |
|  | Exomiser | 55.2 | 81.0 | 93.1 | 96.6 | 96.6 | 98.3 |
|  | PhenoSV | 43.1 | 56.9 | 60.3 | 67.2 | 77.6 | 91.4 |
|  | SvAnna | 0.0 | 0.0 | 0.0 | 0.0 | 0.0 | 0.0 |
| 1–10 kb (*n* = 157) | CoNVict | 71.3 | 85.4 | 89.8 | 91.1 | 93.6 | 93.6 |
|  | Exomiser | 58.6 | 73.2 | 79.0 | 87.3 | 91.7 | 96.2 |
|  | PhenoSV | 38.2 | 55.4 | 59.2 | 63.1 | 72.0 | 84.7 |
|  | SvAnna | 0.0 | 0.6 | 1.3 | 1.9 | 7.0 | 17.8 |
| 10–100 kb (*n* = 74) | CoNVict | 81.1 | 89.2 | 90.5 | 91.9 | 93.2 | 93.2 |
|  | Exomiser | 68.9 | 89.2 | 94.6 | 95.9 | 95.9 | 97.3 |
|  | PhenoSV | 24.3 | 39.2 | 51.4 | 62.2 | 73.0 | 90.5 |
|  | SvAnna | 0.0 | 14.9 | 21.6 | 31.1 | 55.4 | 71.6 |
| 100 kb–1 Mb (*n* = 64) | CoNVict | 82.8 | 96.9 | 100.0 | 100.0 | 100.0 | 100.0 |
|  | Exomiser | 71.9 | 92.2 | 98.4 | 100.0 | 100.0 | 100.0 |
|  | PhenoSV | 26.6 | 51.6 | 60.9 | 70.3 | 78.1 | 89.1 |
|  | SvAnna | 17.2 | 35.9 | 42.2 | 54.7 | 84.4 | 96.9 |
| >1 Mb (*n* = 44) | CoNVict | 79.5 | 93.2 | 97.7 | 97.7 | 100.0 | 100.0 |
|  | Exomiser | 75.0 | 86.4 | 95.5 | 97.7 | 100.0 | 100.0 |
|  | PhenoSV | 65.9 | 81.8 | 84.1 | 88.6 | 90.9 | 97.7 |
|  | SvAnna | 54.5 | 77.3 | 81.8 | 88.6 | 95.5 | 97.7 |

**Table S3.** Top-*K* recall by CNV mechanism. Top-1, Top-5, Top-10, Top-16, Top-32, and Top-64 recall (%) for each of four prioritization tools (`CoNVict`, Exomiser, PhenoSV, SvAnna), stratified by the structural mechanism of the diagnostic CNV: Intragenic (*n* = 203), Whole Gene deletion or duplication (*n* = 24), Gene Fusion via deletion (DEL, *n* = 112), Multi-gene amplification via duplication (DUP, *n* = 44), and Intergenic (*n* = 14). Each sub-table reports results for a single prioritization tool.

| (a) CoNVict |  |  |  |  |  |  |  |
| --- | --- | --- | --- | --- | --- | --- | --- |
| Mechanism | $n$ | Top-1 | Top-5 | Top-10 | Top-16 | Top-32 | Top-64 |
| Intragenic | 203 | 70.4 | 89.2 | 93.6 | 94.6 | 96.6 | 97.0 |
| Whole Gene | 24 | 87.5 | 95.8 | 95.8 | 95.8 | 100.0 | 100.0 |
| Gene Fusion (DEL) | 112 | 84.8 | 94.6 | 96.4 | 96.4 | 97.3 | 97.3 |
| Multi-gene (DUP) | 44 | 75.0 | 93.2 | 100.0 | 100.0 | 100.0 | 100.0 |
| Intergenic | 14 | 21.4 | 21.4 | 28.6 | 35.7 | 35.7 | 35.7 |
| (b) Exomiser |  |  |  |  |  |  |  |
| Intragenic | 203 | 59.6 | 76.8 | 85.2 | 92.6 | 95.1 | 98.0 |
| Whole Gene | 24 | 70.8 | 87.5 | 95.8 | 100.0 | 100.0 | 100.0 |
| Gene Fusion (DEL) | 112 | 77.7 | 95.5 | 98.2 | 98.2 | 99.1 | 100.0 |
| Multi-gene (DUP) | 44 | 65.9 | 88.6 | 97.7 | 97.7 | 97.7 | 97.7 |
| Intergenic | 14 | 0.0 | 14.3 | 28.6 | 42.9 | 50.0 | 64.3 |
| (c) PhenoSV |  |  |  |  |  |  |  |
| Intragenic | 203 | 40.9 | 58.1 | 62.1 | 67.0 | 76.8 | 88.7 |
| Whole Gene | 24 | 25.0 | 41.7 | 50.0 | 66.7 | 75.0 | 91.7 |
| Gene Fusion (DEL) | 112 | 31.2 | 50.0 | 60.7 | 69.6 | 76.8 | 92.0 |
| Multi-gene (DUP) | 44 | 56.8 | 77.3 | 81.8 | 86.4 | 86.4 | 93.2 |
| Intergenic | 14 | 0.0 | 0.0 | 0.0 | 7.1 | 28.6 | 28.6 |
| (d) SvAnna |  |  |  |  |  |  |  |
| Intragenic | 203 | 0.0 | 0.5 | 0.5 | 3.0 | 22.7 | 47.8 |
| Whole Gene | 24 | 0.0 | 0.0 | 0.0 | 4.2 | 29.2 | 45.8 |
| Gene Fusion (DEL) | 112 | 21.4 | 42.0 | 50.9 | 70.5 | 88.4 | 92.9 |
| Multi-gene (DUP) | 44 | 25.0 | 47.7 | 52.3 | 61.4 | 79.5 | 81.8 |
| Intergenic | 14 | 0.0 | 0.0 | 0.0 | 0.0 | 0.0 | 0.0 |

**Table S4.** Top-*K* recall by Sequence Ontology (SO) consequence type. Top-1, Top-5, Top-10, Top-16, Top-32, and Top-64 recall (%) for each of four prioritization tools (`CoNVict`, Exomiser, PhenoSV, SvAnna), stratified by the predicted SO consequence of the diagnostic CNV: Transcript-level variant (*n* = 24), Exon Variant (*n* = 156), Gene Fusion (*n* = 110), Multi-gene Amplification (*n* = 44), Intron Variant (*n* = 49), and Intergenic (*n* = 14). Each sub-table reports results for a single prioritization tool.

| (a) CoNVict |  |  |  |  |  |  |  |
| --- | --- | --- | --- | --- | --- | --- | --- |
| Consequence | $n$ | Top-1 | Top-5 | Top-10 | Top-16 | Top-32 | Top-64 |
| Transcript-level | 24 | 87.5 | 95.8 | 95.8 | 95.8 | 100.0 | 100.0 |
| Exon Variant | 156 | 76.3 | 93.6 | 96.2 | 96.8 | 98.1 | 98.1 |
| Gene Fusion | 110 | 85.5 | 95.5 | 97.3 | 97.3 | 98.2 | 98.2 |
| Multi-gene Amp. | 44 | 75.0 | 93.2 | 100.0 | 100.0 | 100.0 | 100.0 |
| Intron Variant | 49 | 51.0 | 73.5 | 83.7 | 85.7 | 89.8 | 91.8 |
| Intergenic | 14 | 21.4 | 21.4 | 28.6 | 35.7 | 35.7 | 35.7 |
| (b) Exomiser |  |  |  |  |  |  |  |
| Transcript-level | 24 | 70.8 | 87.5 | 95.8 | 100.0 | 100.0 | 100.0 |
| Exon Variant | 156 | 73.1 | 91.0 | 95.5 | 98.7 | 100.0 | 100.0 |
| Gene Fusion | 110 | 79.1 | 97.3 | 100.0 | 100.0 | 100.0 | 100.0 |
| Multi-gene Amp. | 44 | 65.9 | 88.6 | 97.7 | 97.7 | 97.7 | 97.7 |
| Intron Variant | 49 | 14.3 | 28.6 | 49.0 | 69.4 | 77.6 | 91.8 |
| Intergenic | 14 | 0.0 | 14.3 | 28.6 | 42.9 | 50.0 | 64.3 |
| (c) PhenoSV |  |  |  |  |  |  |  |
| Transcript-level | 24 | 25.0 | 41.7 | 50.0 | 66.7 | 75.0 | 91.7 |
| Exon Variant | 156 | 52.6 | 72.4 | 76.3 | 79.5 | 85.3 | 94.2 |
| Gene Fusion | 110 | 31.8 | 50.9 | 61.8 | 70.0 | 77.3 | 92.7 |
| Multi-gene Amp. | 44 | 56.8 | 77.3 | 81.8 | 86.4 | 86.4 | 93.2 |
| Intron Variant | 49 | 2.0 | 10.2 | 14.3 | 26.5 | 49.0 | 69.4 |
| Intergenic | 14 | 0.0 | 0.0 | 0.0 | 7.1 | 28.6 | 28.6 |
| (d) SvAnna |  |  |  |  |  |  |  |
| Transcript-level | 24 | 0.0 | 0.0 | 0.0 | 4.2 | 29.2 | 45.8 |
| Exon Variant | 156 | 0.0 | 0.6 | 0.6 | 3.8 | 29.5 | 62.2 |
| Gene Fusion | 110 | 21.8 | 42.7 | 51.8 | 71.8 | 90.0 | 94.5 |
| Multi-gene Amp. | 44 | 25.0 | 47.7 | 52.3 | 61.4 | 79.5 | 81.8 |
| Intron Variant | 49 | 0.0 | 0.0 | 0.0 | 0.0 | 0.0 | 0.0 |
| Intergenic | 14 | 0.0 | 0.0 | 0.0 | 0.0 | 0.0 | 0.0 |

**Figure S7.**
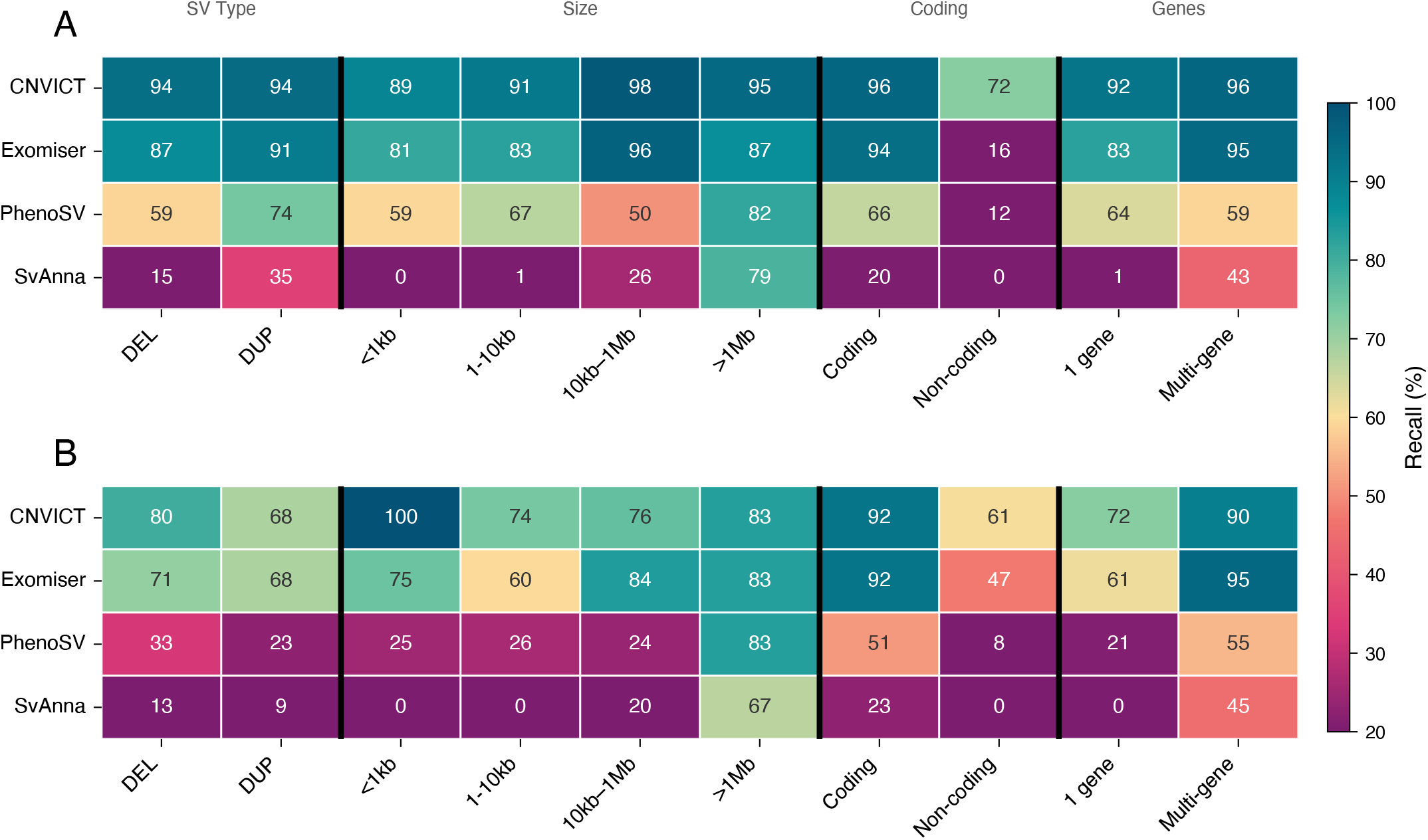
Heatmap of Top-*K* recall across detailed stratification of CNV’s. **(A)** WES cohort, Top-5 recall (%). **(B)** WGS cohort, Top-10 recall (%). Rows represent the four prioritization tools (`CoNVict`, Exomiser, PhenoSV, SvAnna); columns represent clinical strata grouped into four categories separated by black vertical lines: SV Type (DEL, DUP), Size (*<*1 kb, 1–10 kb, 10 kb–1 Mb, *>*1 Mb), Coding status (Coding, Non-coding), and Gene count (1 gene, Multi-gene). Color intensity corresponds to recall percentage, mapped through a diverging purple–teal colormap (purple = low, teal = high; range 20–100%). Numeric recall values are annotated within each cell. This figure differs from the main text heatmap by separating the *<*1 kb size category and combining multi-gene categories (1–3 genes and *>*3 genes) into a single Multi-gene stratum. A shared colorbar is displayed on the right. Group-level category labels (SV Type, Size, Coding, Genes) are displayed above the top panel.

### Performance by Diagnostic Context

**Figure S8.**
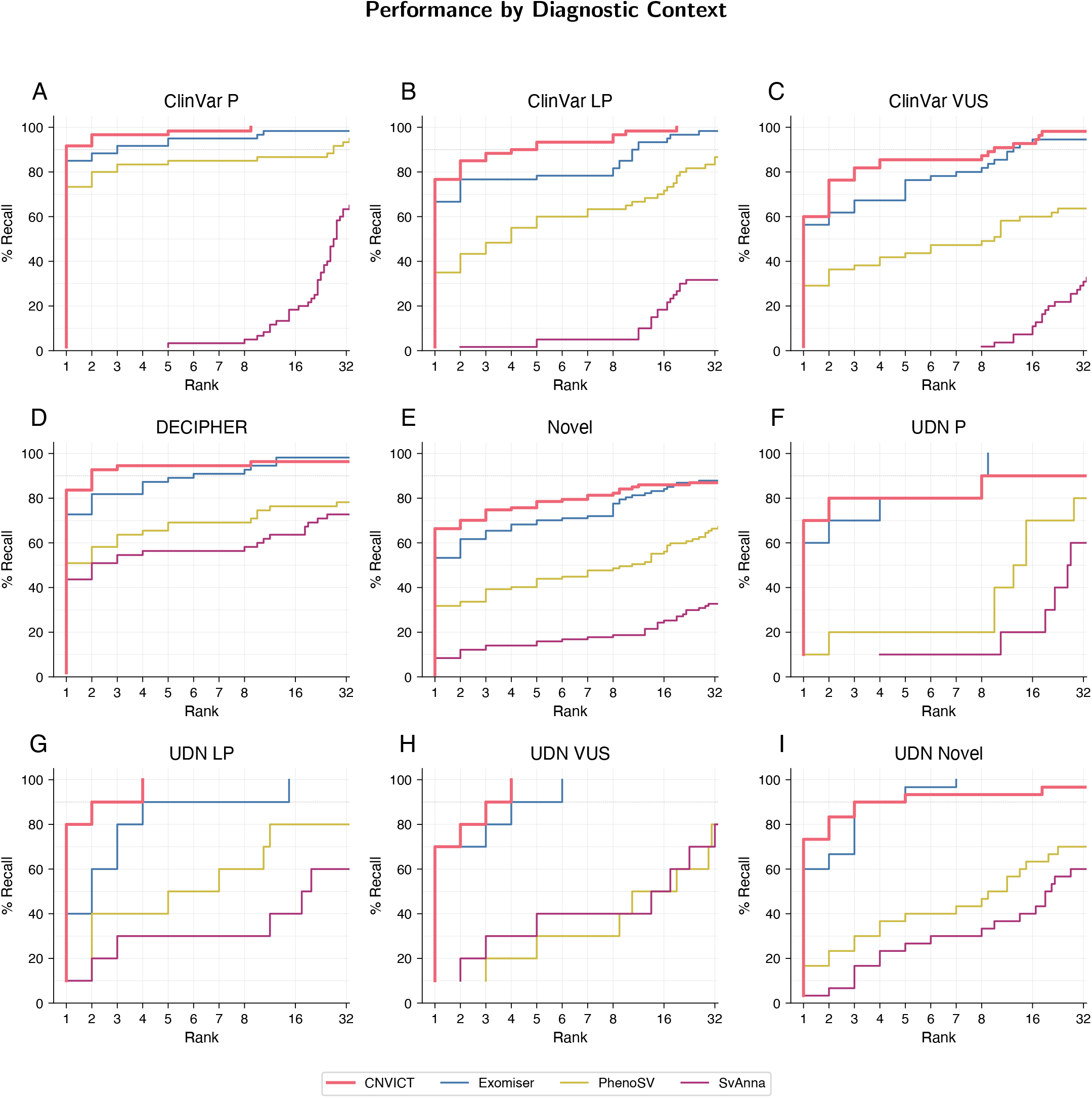
Cumulative distribution of diagnostic CNV ranking stratified by diagnostic CNV source. Each panel displays the CDF of diagnostic CNV rank for all four tools within a specific CNV source category: **(A)** ClinVar Pathogenic (P). **(B)** ClinVar Likely Pathogenic (LP). **(C)** ClinVar Variant of Uncertain Significance (VUS). **(D)** DECIPHER. **(E)** Novel. **(F)** UDN Pathogenic (P). **(G)** UDN Likely Pathogenic (LP). **(H)** UDN Variant of Uncertain Significance (VUS). **(I)** UDN Novel. The piecewise-linear rank scale is used throughout. The grey dotted line marks 90% recall. Panels are arranged in a 3*×*3 grid. A shared legend is provided at the bottom of the figure. See Supplementary Table S5 for Top-1/5/10/16/32/64 recall per tool and source.

**Table S5.** Top-*K* recall by CNV source. Top-1, Top-5, Top-10, Top-16, Top-32, and Top-64 recall (%) for each of four prioritization tools (`CoNVict`, Exomiser, PhenoSV, SvAnna), stratified by nine diagnostic CNV sources: ClinVar Pathogenic (P, *n* = 60), ClinVar Likely Pathogenic (LP, *n* = 60), ClinVar Variant of Uncertain Significance (VUS, *n* = 55), DECIPHER (*n* = 55), Novel (*n* = 107), UDN Pathogenic (P, *n* = 10), UDN Likely Pathogenic (LP, *n* = 10), UDN VUS (*n* = 10), and UDN Novel (*n* = 30). Accompanies Supplementary Figure S4.

| Source | Tool | Top-1 | Top-5 | Top-10 | Top-16 | Top-32 | Top-64 |
| --- | --- | --- | --- | --- | --- | --- | --- |
| ClinVar P (*n* = 60) | CoNVict | **91.7** | **98.3** | **100.0** | **100.0** | **100.0** | **100.0** |
|  | Exomiser | 85.0 | 95.0 | 96.7 | 98.3 | 98.3 | **100.0** |
|  | PhenoSV | 73.3 | 85.0 | 86.7 | 88.3 | 93.3 | 98.3 |
|  | SvAnna | 0.0 | 3.3 | 6.7 | 21.7 | 55.0 | 73.3 |
| ClinVar LP (*n* = 60) | CoNVict | **76.7** | **93.3** | **98.3** | 98.3 | **100.0** | **100.0** |
|  | Exomiser | 66.7 | 78.3 | 85.0 | **96.7** | 98.3 | 98.3 |
|  | PhenoSV | 35.0 | 60.0 | 65.0 | 71.7 | 80.0 | 93.3 |
|  | SvAnna | 0.0 | 5.0 | 5.0 | 11.7 | 33.3 | 56.7 |
| ClinVar VUS (*n* = 55) | CoNVict | **60.0** | **85.5** | **90.9** | **92.7** | **98.2** | **98.2** |
|  | Exomiser | 56.4 | 76.4 | 85.5 | 90.9 | 94.5 | **98.2** |
|  | PhenoSV | 29.1 | 43.6 | 50.9 | 58.2 | 70.9 | 90.9 |
|  | SvAnna | 0.0 | 0.0 | 3.6 | 14.5 | 38.2 | 56.4 |
| DECIPHER (*n* = 55) | CoNVict | **83.6** | **94.5** | 96.4 | 96.4 | 96.4 | 96.4 |
|  | Exomiser | 72.7 | 89.1 | 94.5 | **98.2** | **98.2** | **98.2** |
|  | PhenoSV | 50.9 | 69.1 | 74.5 | 76.4 | 78.2 | 85.5 |
|  | SvAnna | 43.6 | 56.4 | 60.0 | 63.6 | 72.7 | 83.6 |
| Novel (*n* = 107) | CoNVict | **66.4** | **78.5** | 84.1 | **86.0** | 86.9 | 87.9 |
|  | Exomiser | 53.3 | 70.1 | **80.4** | 84.1 | **87.9** | **93.5** |
|  | PhenoSV | 31.8 | 43.9 | 49.5 | 56.1 | 66.4 | 76.6 |
|  | SvAnna | 8.4 | 15.9 | 18.7 | 25.2 | 32.7 | 43.0 |
| UDN P (*n* = 10) | CoNVict | **70.0** | **80.0** | 90.0 | 90.0 | 90.0 | 90.0 |
|  | Exomiser | 60.0 | **80.0** | **100.0** | **100.0** | **100.0** | **100.0** |
|  | PhenoSV | 10.0 | 20.0 | 40.0 | 60.0 | 80.0 | **100.0** |
|  | SvAnna | 0.0 | 10.0 | 10.0 | 20.0 | 40.0 | 50.0 |
| UDN LP (*n* = 10) | CoNVict | **80.0** | **100.0** | **100.0** | **100.0** | **100.0** | **100.0** |
|  | Exomiser | 40.0 | 90.0 | 90.0 | **100.0** | **100.0** | **100.0** |
|  | PhenoSV | 0.0 | 50.0 | 60.0 | 60.0 | 70.0 | 90.0 |
|  | SvAnna | 10.0 | 30.0 | 30.0 | 30.0 | 50.0 | 80.0 |
| UDN VUS (*n* = 10) | CoNVict | **70.0** | **100.0** | **100.0** | **100.0** | **100.0** | **100.0** |
|  | Exomiser | **70.0** | 90.0 | **100.0** | **100.0** | **100.0** | **100.0** |
|  | PhenoSV | 0.0 | 30.0 | 40.0 | 60.0 | 60.0 | 80.0 |
|  | SvAnna | 0.0 | 40.0 | 40.0 | 50.0 | 80.0 | 90.0 |
| UDN Novel (*n* = 30) | CoNVict | **73.3** | 93.3 | 93.3 | 93.3 | **100.0** | **100.0** |
|  | Exomiser | 60.0 | **96.7** | **100.0** | **100.0** | **100.0** | **100.0** |
|  | PhenoSV | 16.7 | 40.0 | 50.0 | 66.7 | 76.7 | 93.3 |
|  | SvAnna | 3.3 | 26.7 | 36.7 | 43.3 | 70.0 | 80.0 |

**Figure S9.**
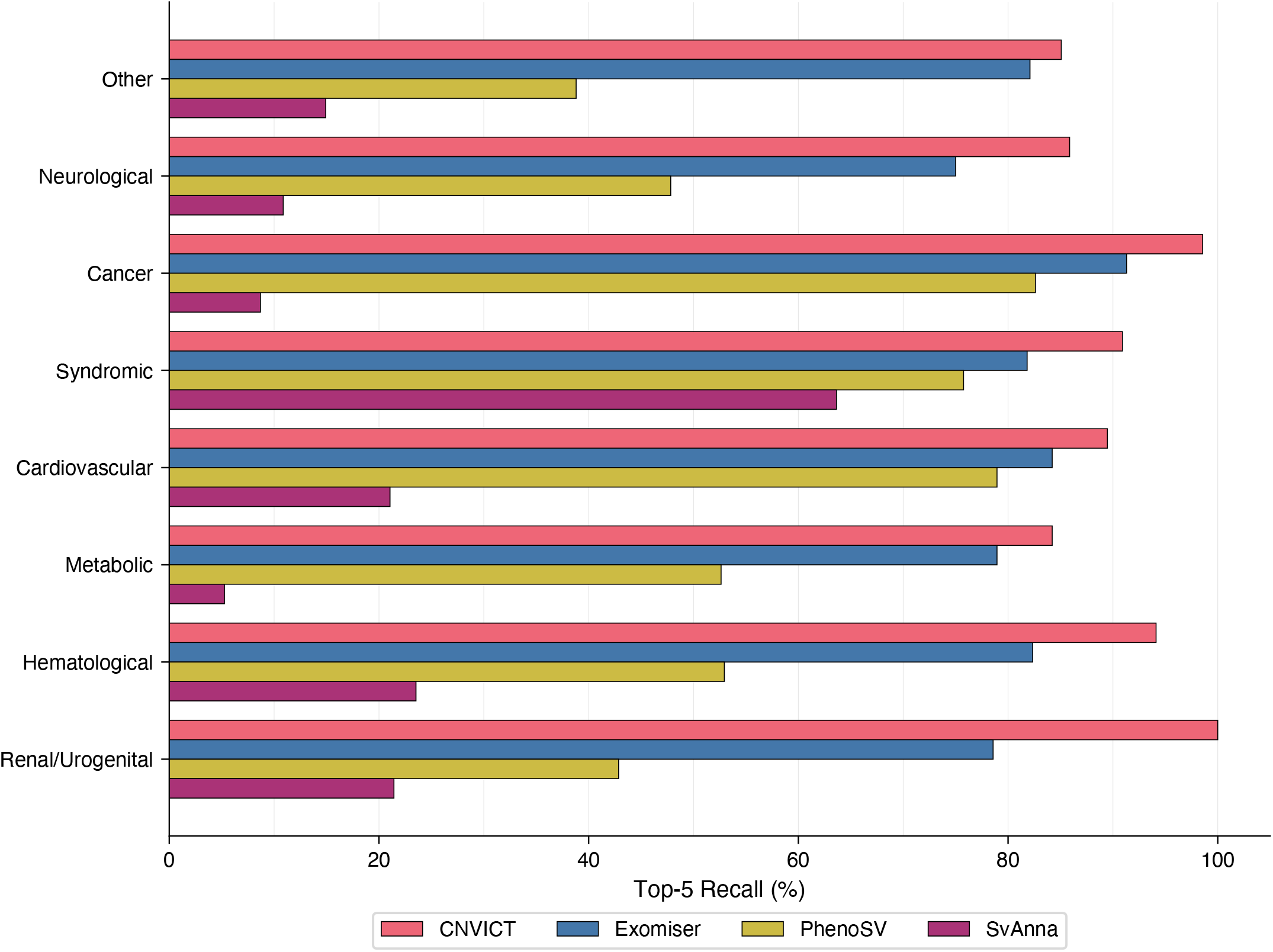
Top-5 recall by disease system across prioritization tools. Horizontal grouped bars show the percentage of diagnostic CNVs ranked within the Top-5 for each prioritization tool (`CoNVict`, Exomiser, PhenoSV, SvAnna), stratified by the disease system of the underlying disorder. Disease systems are sorted in descending order by sample size. Only systems with at least 5 patients are included for statistical reliability. Thin horizontal grid lines are drawn at 10% intervals. A shared legend is provided below the plot. See Supplementary Table S6 for Top-1/5/10/16/32/64 recall per tool and disease system.

**Table S6.** Top-*K* recall by disease system. Top-1, Top-5, Top-10, Top-16, Top-32, and Top-64 recall (%) for each of four prioritization tools (`CoNVict`, Exomiser, PhenoSV, SvAnna), stratified by eight disease systems: Other (*n* = 134), Neurological (*n* = 92), Cancer (*n* = 69), Syndromic (*n* = 33), Cardiovascular (*n* = 19), Metabolic (*n* = 19), Hematological (*n* = 17), and Renal/Urogenital (*n* = 14). Only systems with *n* ≥ 5 are included. Accompanies Supplementary Figure S5.

| Disease System | Tool | Top-1 | Top-5 | Top-10 | Top-16 | Top-32 | Top-64 |
| --- | --- | --- | --- | --- | --- | --- | --- |
| Other ($n = 134$) | CoNVict | **67.2** | **85.1** | **90.3** | 91.0 | 93.3 | 93.3 |
|  | Exomiser | 57.5 | 82.1 | 89.6 | **94.0** | **94.0** | **96.3** |
|  | PhenoSV | 16.4 | 38.8 | 47.0 | 58.2 | 67.2 | 85.1 |
|  | SvAnna | 2.2 | 14.9 | 19.4 | 26.9 | 44.0 | 61.2 |
| Neurological ($n = 92$) | CoNVict | **65.2** | **85.9** | **90.2** | **91.3** | 92.4 | 93.5 |
|  | Exomiser | 58.7 | 75.0 | 83.7 | 89.1 | **93.5** | **97.8** |
|  | PhenoSV | 34.8 | 47.8 | 52.2 | 56.5 | 67.4 | 84.8 |
|  | SvAnna | 8.7 | 10.9 | 15.2 | 25.0 | 34.8 | 52.2 |
| Cancer ($n = 69$) | CoNVict | **94.2** | **98.6** | **98.6** | **98.6** | 98.6 | 98.6 |
|  | Exomiser | 81.2 | 91.3 | 95.7 | 97.1 | **100.0** | **100.0** |
|  | PhenoSV | 66.7 | 82.6 | 85.5 | 87.0 | 94.2 | 97.1 |
|  | SvAnna | 4.3 | 8.7 | 10.1 | 20.3 | 63.8 | 75.4 |
| Syndromic ($n = 33$) | CoNVict | **75.8** | **90.9** | **93.9** | 93.9 | 93.9 | 93.9 |
|  | Exomiser | 66.7 | 81.8 | 90.9 | **97.0** | **97.0** | **97.0** |
|  | PhenoSV | 51.5 | 75.8 | 81.8 | 87.9 | 87.9 | 87.9 |
|  | SvAnna | 45.5 | 63.6 | 63.6 | 66.7 | 72.7 | 78.8 |
| Cardiovascular ($n = 19$) | CoNVict | **78.9** | **89.5** | **100.0** | **100.0** | **100.0** | **100.0** |
|  | Exomiser | 73.7 | 84.2 | 84.2 | 89.5 | 94.7 | 94.7 |
|  | PhenoSV | 63.2 | 78.9 | 78.9 | 84.2 | 94.7 | 94.7 |
|  | SvAnna | 10.5 | 21.1 | 21.1 | 26.3 | 57.9 | 68.4 |
| Metabolic ($n = 19$) | CoNVict | **78.9** | **84.2** | **89.5** | **94.7** | **100.0** | **100.0** |
|  | Exomiser | 63.2 | 78.9 | **89.5** | 89.5 | 89.5 | 94.7 |
|  | PhenoSV | 42.1 | 52.6 | 52.6 | 63.2 | 68.4 | 84.2 |
|  | SvAnna | 0.0 | 5.3 | 10.5 | 21.1 | 26.3 | 52.6 |
| Hematological ($n = 17$) | CoNVict | **64.7** | **94.1** | 94.1 | 94.1 | **100.0** | **100.0** |
|  | Exomiser | 52.9 | 82.4 | **94.1** | **100.0** | **100.0** | **100.0** |
|  | PhenoSV | 41.2 | 52.9 | 64.7 | 76.5 | 88.2 | 94.1 |
|  | SvAnna | 11.8 | 23.5 | 23.5 | 29.4 | 41.2 | 58.8 |
| Renal/Urogenital ($n = 14$) | CoNVict | **100.0** | **100.0** | **100.0** | **100.0** | **100.0** | **100.0** |
|  | Exomiser | 71.4 | 78.6 | 78.6 | 92.9 | 92.9 | **100.0** |
|  | PhenoSV | 35.7 | 42.9 | 64.3 | 64.3 | 71.4 | 85.7 |
|  | SvAnna | 14.3 | 21.4 | 21.4 | 28.6 | 35.7 | 50.0 |

**Figure S10.**
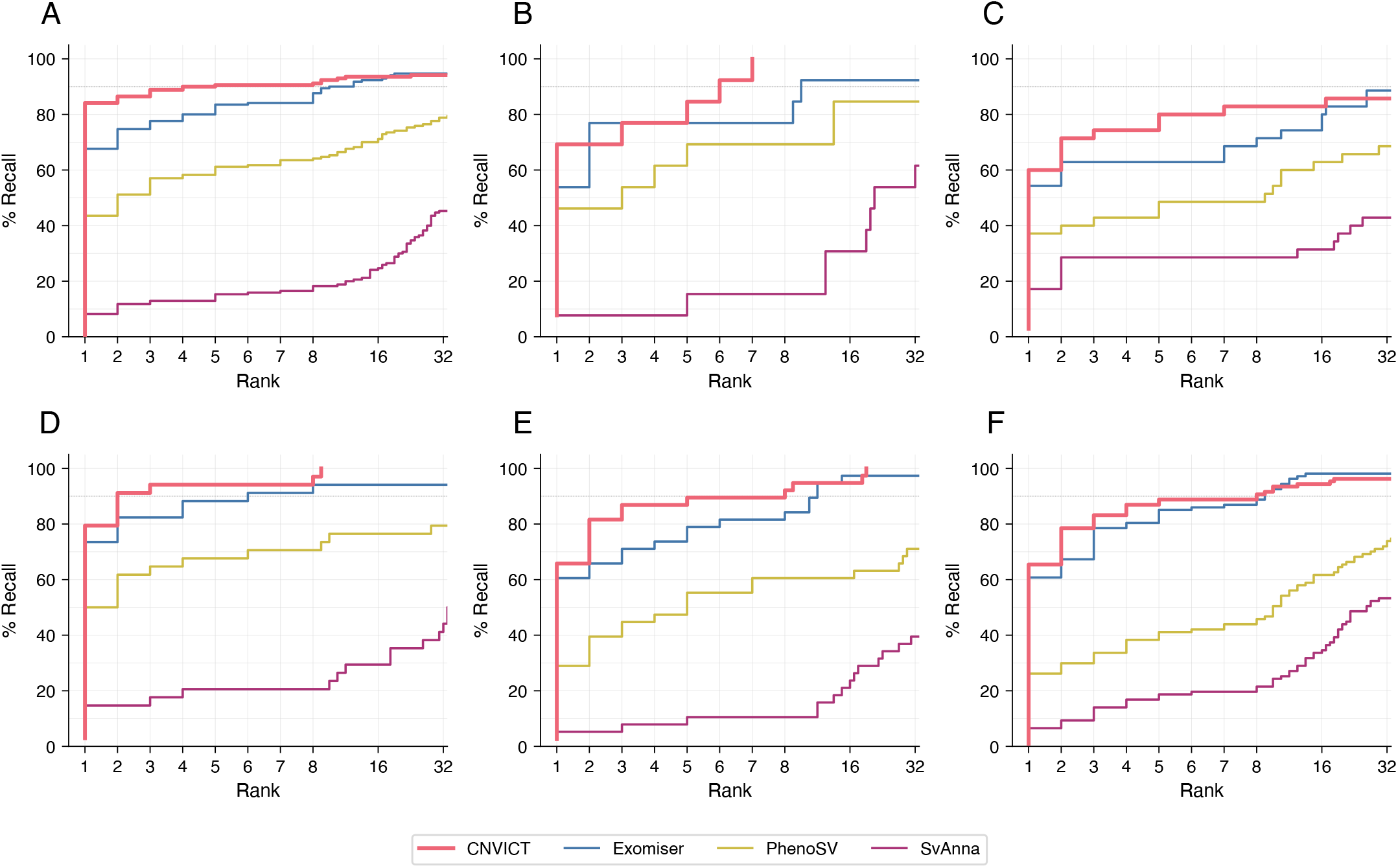
Cumulative distribution of diagnostic CNV ranking stratified by ClinGen dosage sensitivity evidence. Each panel shows the CDF of diagnostic CNV rank for all four tools, stratified by the ClinGen dosage sensitivity score assigned to the gene(s) overlapping the diagnostic CNV. For deletions, the ClinGen Haploinsufficiency (HI) score is used; for duplications, the ClinGen Triplosensitivity (TS) score is used. Panels are arranged in a 2*×*3 grid: **(A)** Sufficient evidence (score 3). **(B)** Some evidence (score 2). **(C)** Little evidence (score 1). **(D)** No evidence (score 0). **(E)** Autosomal recessive phenotype (score 30). None/NA (score not available or not assessed). The piecewise-linear rank scale is used throughout. The grey dotted line marks 90% recall. A shared legend is provided at the bottom of the figure. See Supplementary Table S7 for Top-*K* recall per tool and ClinGen evidence group.

**Table S7.** Top-*K* recall by ClinGen dosage sensitivity group. Top-1, Top-5, Top-10, Top-16, Top-32, and Top-64 recall (%) for each of four prioritization tools (`CoNVict`, Exomiser, PhenoSV, SvAnna), stratified by ClinGen dosage sensitivity score: Sufficient evidence (score 3, *n* = 170), Some evidence (score 2, *n* = 13), Little evidence (score 1, *n* = 35), No evidence (score 0, *n* = 34), Autosomal Recessive (AR) phenotype (score 30, *n* = 38), and None/NA (score not available or not assessed, *n* = 107). For deletions the Haploinsufficiency (HI) score is used; for duplications the Triplosensitivity (TS) score is used. Accompanies Supplementary Figure S6.

| Dosage Score | Tool | Top-1 | Top-5 | Top-10 | Top-16 | Top-32 | Top-64 |
| --- | --- | --- | --- | --- | --- | --- | --- |
| Sufficient, 3<br>(*n* = 170) | CoNVict | 84.1 | 90.6 | 92.4 | 93.5 | 94.1 | 94.7 |
|  | Exomiser | 67.6 | 83.5 | 90.0 | 92.4 | 94.7 | 97.6 |
|  | PhenoSV | 43.5 | 61.2 | 65.3 | 68.8 | 77.1 | 90.6 |
|  | SvAnna | 8.2 | 15.3 | 18.2 | 28.8 | 50.0 | 67.6 |
| Some, 2 (*n* = 13) | CoNVict | 69.2 | 84.6 | 100.0 | 100.0 | 100.0 | 100.0 |
|  | Exomiser | 53.8 | 76.9 | 92.3 | 100.0 | 100.0 | 100.0 |
|  | PhenoSV | 46.2 | 69.2 | 69.2 | 76.9 | 84.6 | 100.0 |
|  | SvAnna | 7.7 | 15.4 | 15.4 | 23.1 | 38.5 | 53.8 |
| Little, 1 (*n* = 35) | CoNVict | 60.0 | 80.0 | 82.9 | 85.7 | 88.6 | 88.6 |
|  | Exomiser | 54.3 | 62.9 | 71.4 | 80.0 | 85.7 | 94.3 |
|  | PhenoSV | 37.1 | 48.6 | 54.3 | 60.0 | 68.6 | 82.9 |
|  | SvAnna | 17.1 | 28.6 | 28.6 | 34.3 | 48.6 | 62.9 |
| No evidence, 0<br>(*n* = 34) | CoNVict | 79.4 | 94.1 | 100.0 | 100.0 | 100.0 | 100.0 |
|  | Exomiser | 73.5 | 88.2 | 94.1 | 100.0 | 100.0 | 100.0 |
|  | PhenoSV | 50.0 | 67.6 | 76.5 | 85.3 | 91.2 | 97.1 |
|  | SvAnna | 14.7 | 20.6 | 23.5 | 29.4 | 52.9 | 64.7 |
| AR phenotype, 30<br>(*n* = 38) | CoNVict | 65.8 | 89.5 | 94.7 | 94.7 | 97.4 | 97.4 |
|  | Exomiser | 60.5 | 78.9 | 84.2 | 92.1 | 94.7 | 97.4 |
|  | PhenoSV | 28.9 | 55.3 | 60.5 | 71.1 | 78.9 | 86.8 |
|  | SvAnna | 5.3 | 10.5 | 10.5 | 13.2 | 23.7 | 39.5 |
| None/NA (*n* = 107) | CoNVict | 65.4 | 88.8 | 93.5 | 94.4 | 96.3 | 97.2 |
|  | Exomiser | 60.7 | 85.0 | 92.5 | 95.3 | 96.3 | 98.1 |
|  | PhenoSV | 26.2 | 41.1 | 50.5 | 61.7 | 72.9 | 83.2 |
|  | SvAnna | 6.5 | 18.7 | 24.3 | 30.8 | 49.5 | 62.6 |

**Figure S11.**
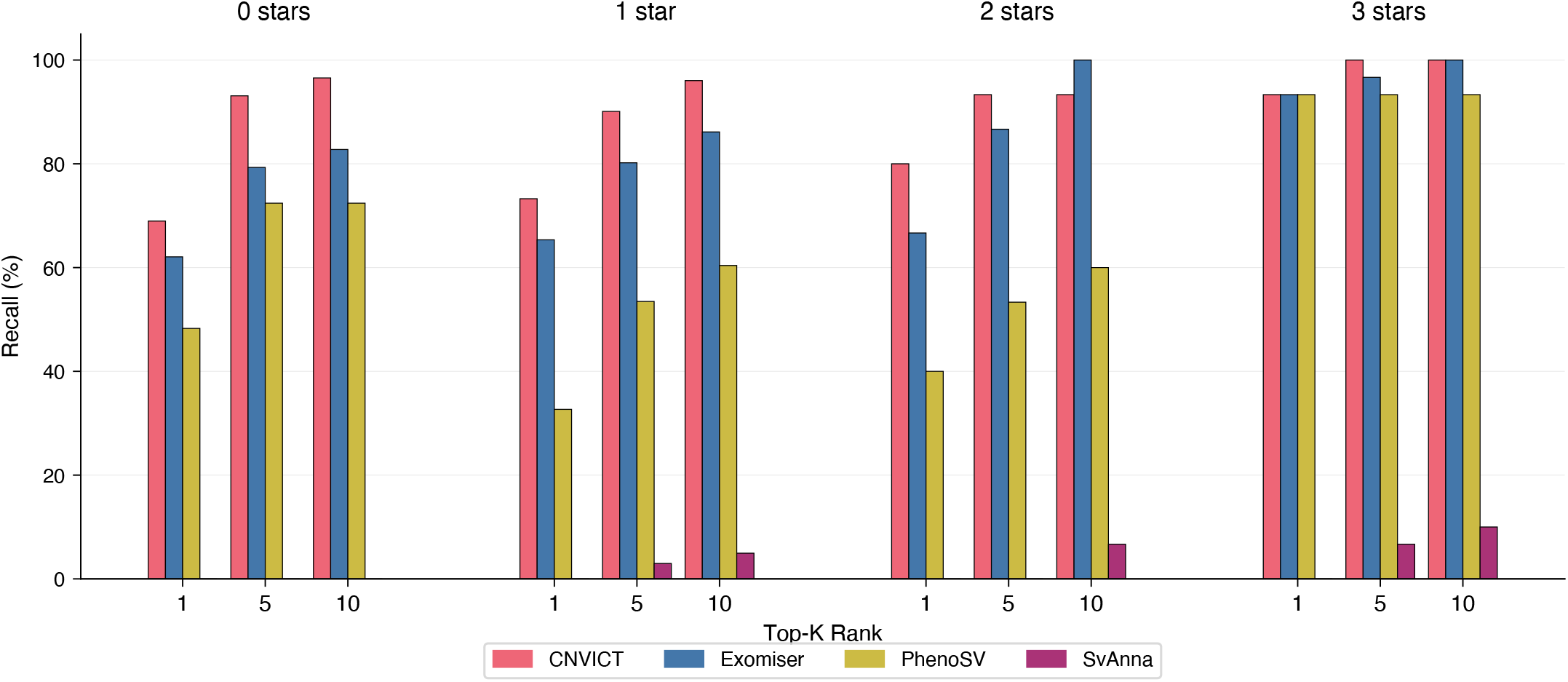
Top-*K* recall by ClinVar review star rating. Grouped bars show recall at Top-1, Top-5, and Top-10 for each prioritization tool (`CoNVict`, Exomiser, PhenoSV, SvAnna), stratified by the ClinVar review star rating of the diagnostic CNV source variant (0 stars, 1 star, 2 stars, 3 stars). Only ClinVar-sourced patients with assigned review stars are included. Within each star rating group, three clusters of bars correspond to the three Top-*K* thresholds, with each cluster containing one bar per tool colored by tool identity. Primary *x*-axis tick labels indicate the Top-*K* threshold; a secondary *x*-axis labels the star rating groups. Higher star ratings indicate stronger evidence support for the ClinVar assertion. Horizontal grid lines are drawn at 20% intervals. A tool-color legend is provided below the plot.

**Figure S12.**
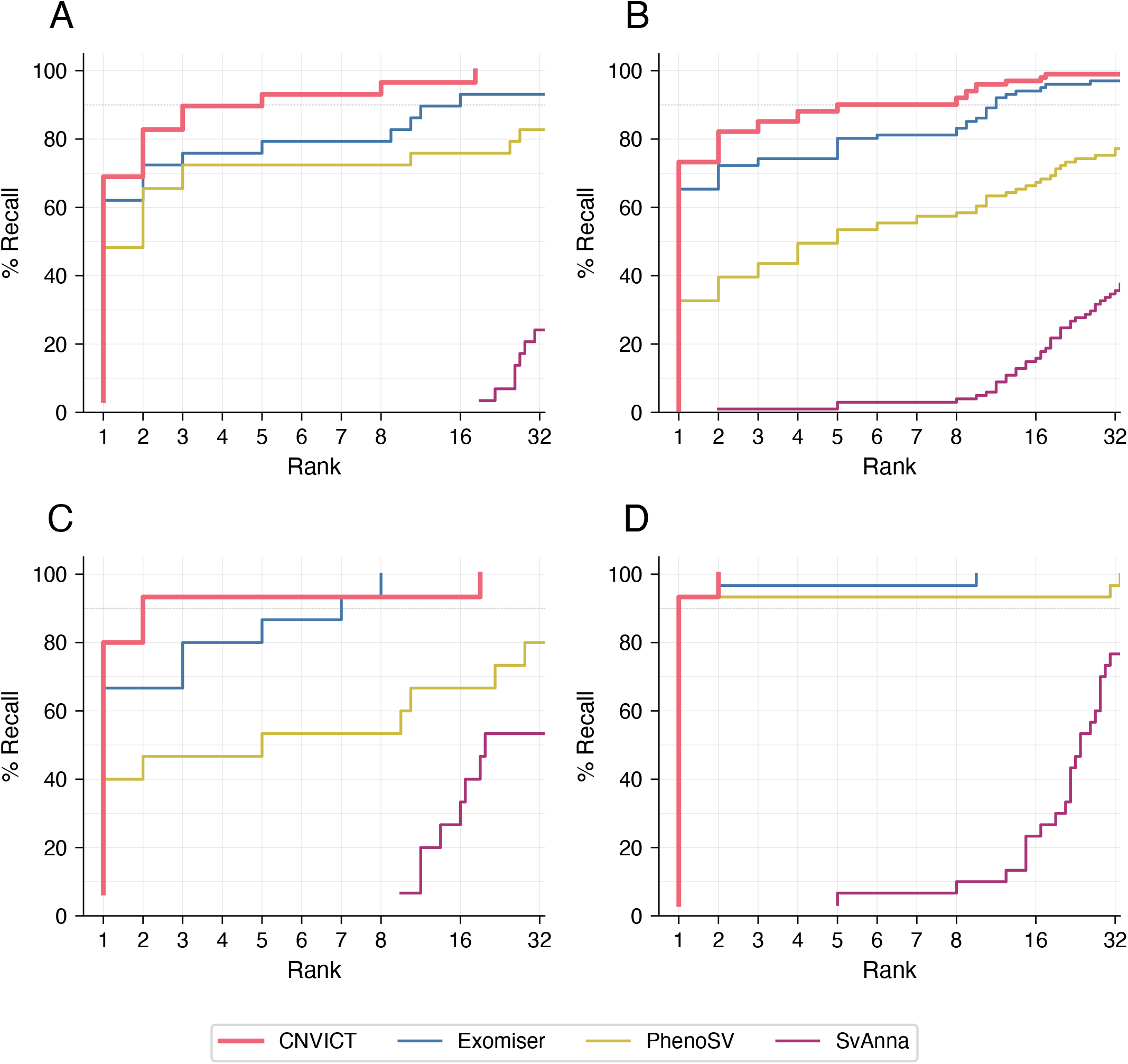
Cumulative distribution of diagnostic CNV ranking stratified by ClinVar review star rating. **(A)** 0 stars. **(B)** 1 star. **(C)** 2 stars. **(D)** 3 stars. Each panel shows the CDF of diagnostic CNV rank for all four tools within a given star rating group. Only ClinVar-sourced patients with assigned review stars are included. The piecewise-linear rank scale is used throughout, emphasizing performance at the top of the ranked list. The grey dotted line marks 90% recall. Panels are arranged in a 2*×*2 grid without subplot titles; panel identity is indicated by the **(A)**–**(D)** labels. A shared legend is provided at the bottom of the figure.

**Table S8.** Top-*K* recall for Variants of Uncertain Significance (VUS) and AnnotSV ACMG VUS subsets. Top-1, Top-5, Top-10, Top-16, Top-32, and Top-64 recall (%) for each of four prioritization tools (`CoNVict`, Exomiser, PhenoSV, SvAnna) on two variant subsets: All VUS (ClinVar VUS, *n* = 65; includes both ClinVar VUS and UDN VUS patients) and AnnotSV ACMG VUS (VUS plus Novel variants, *n* = 124; representing the most clinically challenging cases that lack definitive pathogenicity classification). Each sub-table reports results for a single prioritization tool.

| (a) CoNVict |  |  |  |  |  |  |  |
| --- | --- | --- | --- | --- | --- | --- | --- |
| Subset | $n$ | Top-1 | Top-5 | Top-10 | Top-16 | Top-32 | Top-64 |
| All VUS | 65 | 61.5 | 87.7 | 92.3 | 93.8 | 98.5 | 98.5 |
| AnnotSV ACMG VUS | 124 | 54.8 | 75.8 | 83.1 | 84.7 | 86.3 | 87.1 |
| (b) Exomiser |  |  |  |  |  |  |  |
| All VUS | 65 | 58.5 | 78.5 | 87.7 | 95.4 | 95.4 | 96.9 |
| AnnotSV ACMG VUS | 124 | 38.7 | 58.1 | 72.6 | 83.9 | 88.7 | 94.4 |
| (c) PhenoSV |  |  |  |  |  |  |  |
| All VUS | 65 | 24.6 | 41.5 | 49.2 | 58.5 | 66.2 | 89.2 |
| AnnotSV ACMG VUS | 124 | 18.5 | 33.1 | 36.3 | 46.0 | 59.7 | 74.2 |
| (d) SvAnna |  |  |  |  |  |  |  |
| All VUS | 65 | 0.0 | 6.2 | 9.2 | 16.9 | 38.5 | 58.5 |
| AnnotSV ACMG VUS | 124 | 0.8 | 1.6 | 1.6 | 4.8 | 10.5 | 30.6 |

**Table S9.** Head-to-head pairwise comparison on VUS and AnnotSV ACMG VUS subsets. For each patient, the tool that assigns a lower (better) rank to the diagnostic CNV wins the comparison. Win, tie, and loss percentages are reported for `CoNVict` versus each competitor (Exomiser, PhenoSV, SvAnna). Results are shown separately for the All VUS subset (*n* = 65; ClinVar VUS and UDN VUS patients) and the AnnotSV ACMG VUS subset (*n* = 124; VUS plus Novel variants).

| Subset | vs | CoNVict Wins (%) | Tie (%) | Competitor Wins (%) |
| --- | --- | --- | --- | --- |
| All VUS ($n = 65$) | Exomiser | 33.8 | 46.2 | 20.0 |
|  | PhenoSV | 67.7 | 24.6 | 7.7 |
|  | SvAnna | 98.5 | 0.0 | 1.5 |
| AnnotSV ACMG VUS ($n = 124$) | Exomiser | 46.8 | 28.2 | 25.0 |
|  | PhenoSV | 68.5 | 20.2 | 11.3 |
|  | SvAnna | 95.2 | 1.6 | 3.2 |

**Table S10.** Head-to-head pairwise comparison on the Novel subset (*n* = 107). These 107 patients have diagnostic CNVs that are not present in any curated database and were derived from literature case reports. For each patient, the tool that assigns a lower (better) rank to the diagnostic CNV wins the comparison. Win, tie, and loss percentages are reported for `CoNVict` versus each competitor (Exomiser, PhenoSV, SvAnna).

| vs | CoNVict Wins (%) | Tie (%) | Competitor Wins (%) |
| --- | --- | --- | --- |
| Exomiser | 33.6 | 43.9 | 22.4 |
| PhenoSV | 57.9 | 29.9 | 12.1 |
| SvAnna | 86.9 | 10.3 | 2.8 |

**Table S11.**
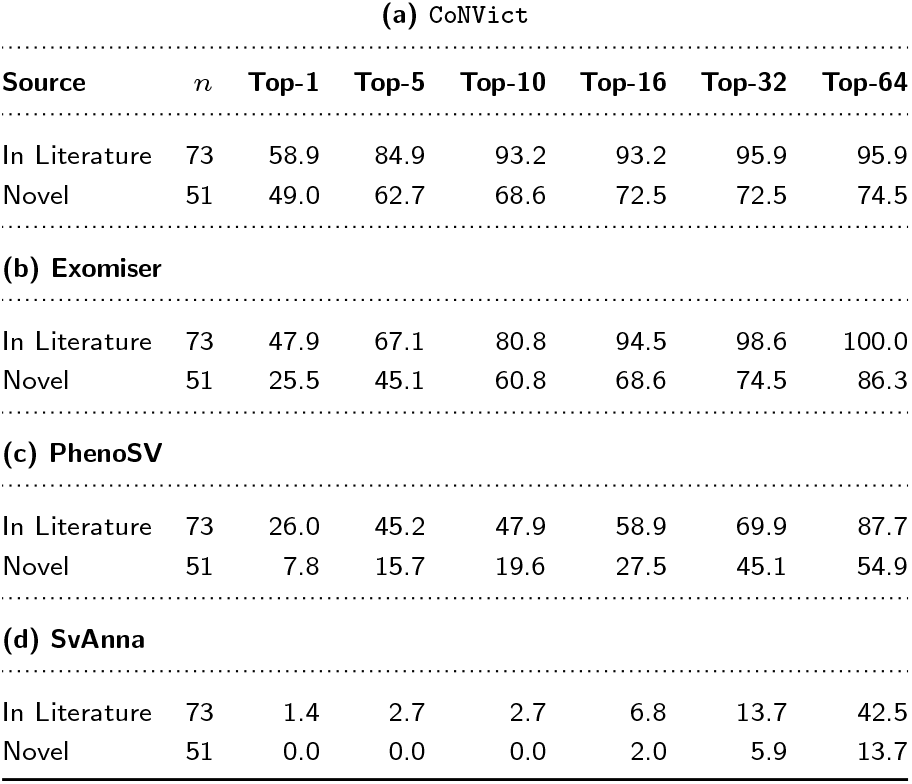
Top-*K* recall for AnnotSV ACMG VUS variants stratified by literature evidence status. Top-1, Top-5, Top-10, Top-16, Top-32, and Top-64 recall (%) for each of four prioritization tools (`CoNVict`, Exomiser, PhenoSV, SvAnna) on the AnnotSV ACMG VUS subset (*n* = 124), split by whether the diagnostic CNV has prior literature support (“In Literature”, *n* = 73; the variant or overlapping gene has been reported in published case reports) or is entirely novel (“Novel”, *n* = 51; no prior literature evidence exists). Each sub-table reports results for a single prioritization tool.

| (a) CoNVict |  |  |  |  |  |  |  |
| --- | --- | --- | --- | --- | --- | --- | --- |
| Source | *n* | Top-1 | Top-5 | Top-10 | Top-16 | Top-32 | Top-64 |
| In Literature | 73 | 58.9 | 84.9 | 93.2 | 93.2 | 95.9 | 95.9 |
| Novel | 51 | 49.0 | 62.7 | 68.6 | 72.5 | 72.5 | 74.5 |
| (b) Exomiser |  |  |  |  |  |  |  |
| In Literature | 73 | 47.9 | 67.1 | 80.8 | 94.5 | 98.6 | 100.0 |
| Novel | 51 | 25.5 | 45.1 | 60.8 | 68.6 | 74.5 | 86.3 |
| (c) PhenoSV |  |  |  |  |  |  |  |
| In Literature | 73 | 26.0 | 45.2 | 47.9 | 58.9 | 69.9 | 87.7 |
| Novel | 51 | 7.8 | 15.7 | 19.6 | 27.5 | 45.1 | 54.9 |
| (d) SvAnna |  |  |  |  |  |  |  |
| In Literature | 73 | 1.4 | 2.7 | 2.7 | 6.8 | 13.7 | 42.5 |
| Novel | 51 | 0.0 | 0.0 | 0.0 | 2.0 | 5.9 | 13.7 |

### UDN Cohort Analysis

**Figure S13.**
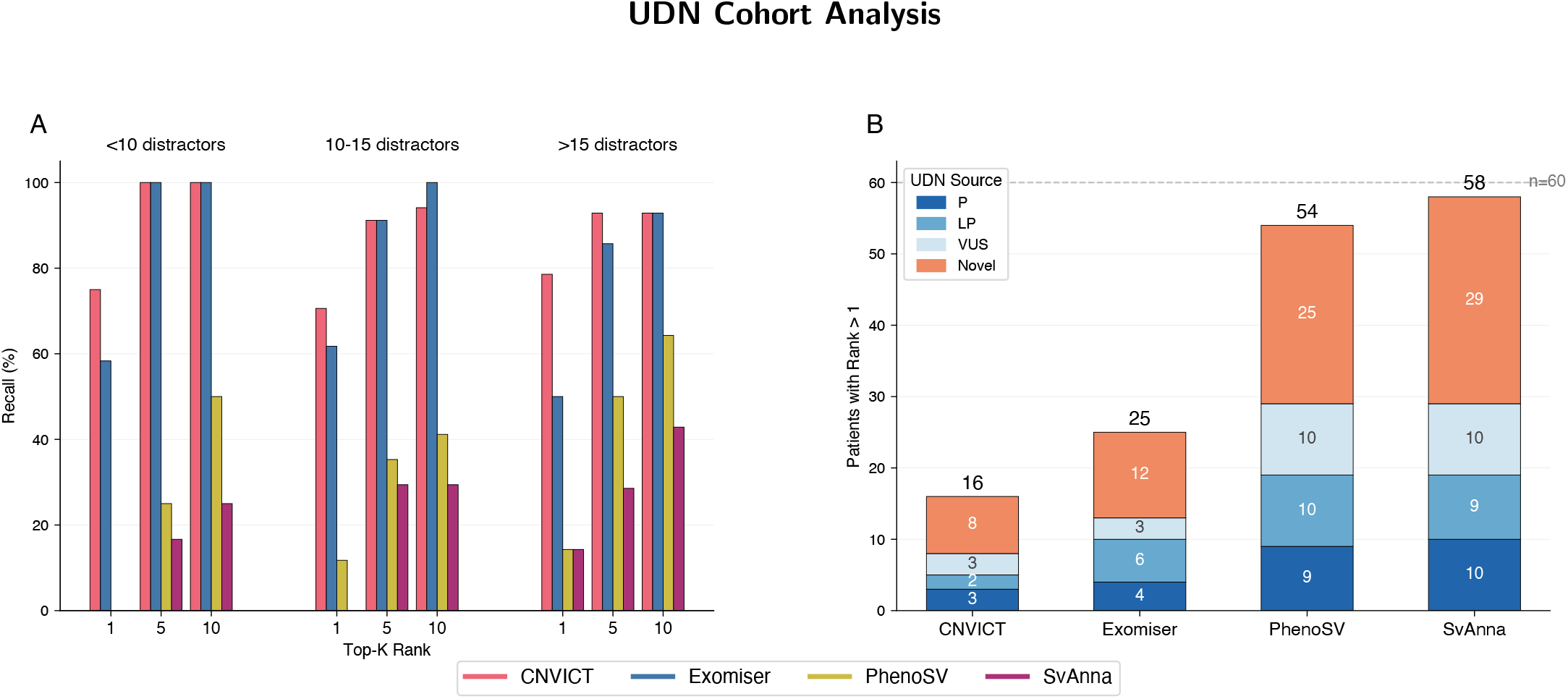
UDN distractor analysis. **(A)** Top-*K* recall by distractor CNV burden. UDN patients are binned by the number of additional clinically reported CNVs (distractors) in their case: *<*10, 10–15, and *>*15. Within each distractor bin, grouped bars display recall at Top-1, Top-5, and Top-10 for four tools: `CoNVict`, Exomiser, PhenoSV, and SvAnna. Primary *x*-axis tick labels indicate the Top-*K* threshold; a secondary *x*-axis labels the distractor burden bins. **(B)** Missed case analysis. Stacked bars show the number of UDN patients for whom each tool failed to rank the diagnostic CNV at rank 1, broken down by diagnostic CNV source: ClinVar Pathogenic (P), ClinVar Likely Pathogenic (LP), ClinVar Variant of Uncertain Significance (VUS), and Novel. Numeric counts are annotated within each segment where space permits, and total missed counts are annotated above each bar. The dashed horizontal line marks the total UDN cohort size (*n* = 60). Novel cases dominate the missed fraction across all tools, reflecting the inherent difficulty of prioritizing previously unreported diagnostic CNVs. A shared tool-color legend is provided at the bottom of the figure.

**Figure S14.**
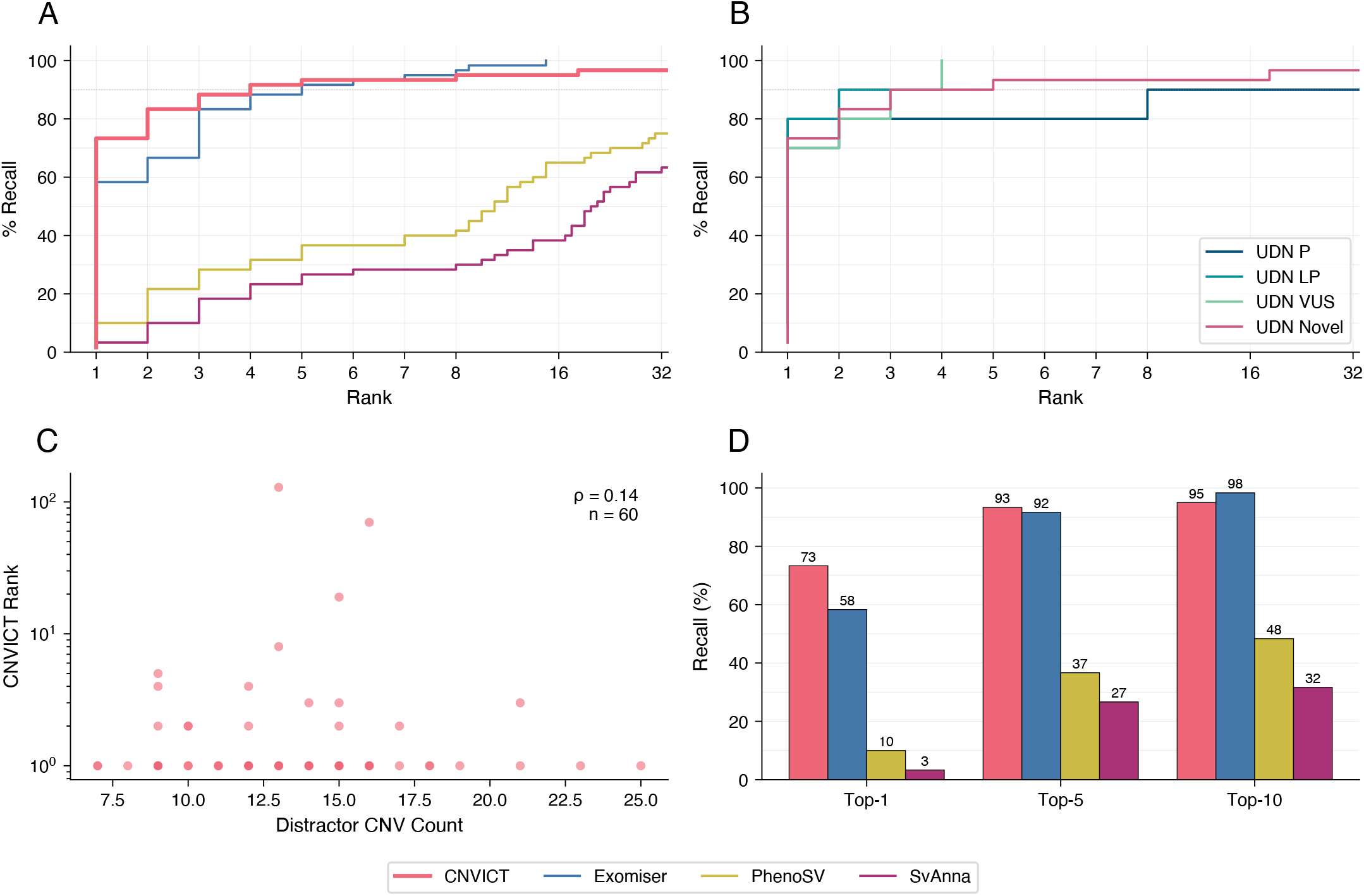
UDN cohort deep-dive analysis. **(A)** CDF of diagnostic CNV rank for all four tools on the UDN subset. **(B)** CDF by UDN sub-source (`CoNVict` only): ClinVar Pathogenic (P), ClinVar Likely Pathogenic (LP), ClinVar Variant of Uncertain Significance (VUS), and Novel diagnostic CNVs. Each sub-source curve uses a distinct color. **(C)** Distractor impact scatter plot showing the relationship between distractor CNV count (*x*-axis) and `CoNVict` rank (*y*-axis, log scale). Spearman correlation coefficient (*ρ*) and sample size are annotated in the upper right corner. **(D)** Top-*K* grouped bar chart (Top-1, Top-5, Top-10) for all four tools on the UDN subset. The piecewise-linear rank scale is used in CDF panels. A shared legend for the four tools is provided at the bottom of the figure.

**Figure S15.**
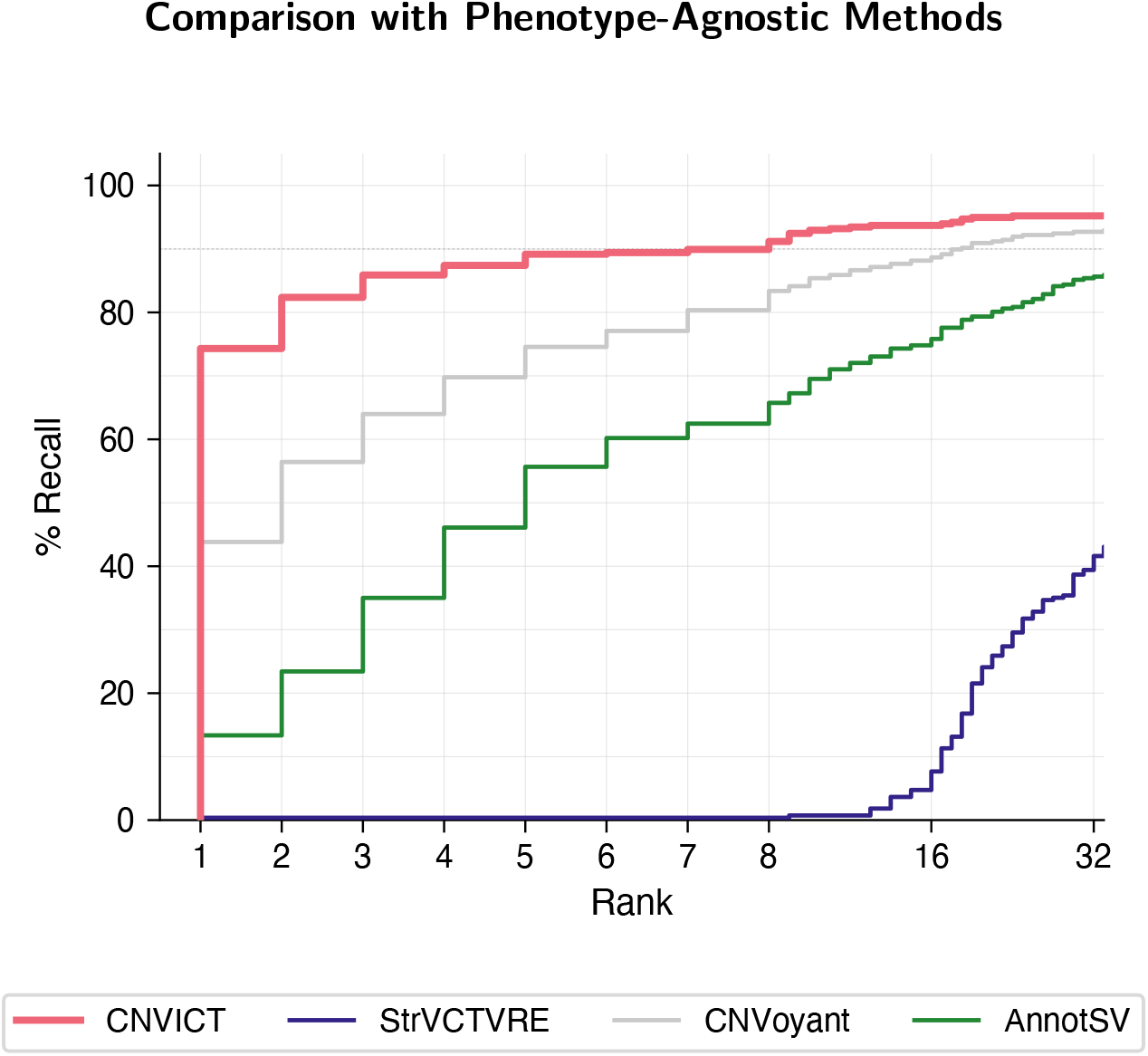
Cumulative recall of phenotype-agnostic CNV scoring tools. Cumulative distribution of diagnostic variant rank across 397 simulated patients for four phenotype-independent tools: `CoNVict`, StrVCTVRE, CNVoyant, and AnnotSV. Rank 1 indicates the diagnostic CNV received the highest score among all variants in a given patient. `CoNVict` ranked the diagnostic variant first in 74.3% of cases (top-3: 85.9%, top-10: 92.9%). CNVoyant achieved 43.8% top-1 recall (top-10: 85.4%). AnnotSV, which assigns ACMG-style classifications rather than continuous pathogenicity scores, reached 13.4% top-1 recall (top-10: 69.5%). StrVCTVRE scores only exonic DEL/DUP variants and could evaluate 274 of 397 patients (top-10: 0.7%). The *x*-axis uses a piecewise-linear scale that expands ranks 1–8 where clinical actionability is highest. The dashed horizontal line marks 90% recall.

**Table S12.** Phenotype-agnostic tool performance summary. Top-*K* recall (%) and median rank for each tool across all 397 simulated patients. StrVCTVRE evaluated 274 patients (exonic diagnostic CNVs only); all other tools scored all 397

| Tool | *n* | Top-1 | Top-3 | Top-5 | Top-10 | Top-16 | Top-32 | Median |
| --- | --- | --- | --- | --- | --- | --- | --- | --- |
| CoNVict | 397 | 74.3 | 85.9 | 89.2 | 92.9 | 93.7 | 95.2 | 1.0 |
| CNVoyant | 397 | 43.8 | 64.0 | 74.6 | 85.4 | 88.7 | 92.7 | 2.0 |
| AnnotSV | 397 | 13.4 | 35.0 | 55.7 | 69.5 | 75.8 | 85.6 | 5.0 |
| StrVCTVRE | 274 | 0.4 | 0.4 | 0.4 | 0.7 | 7.7 | 41.6 | 41.0 |

**Figure S16.**
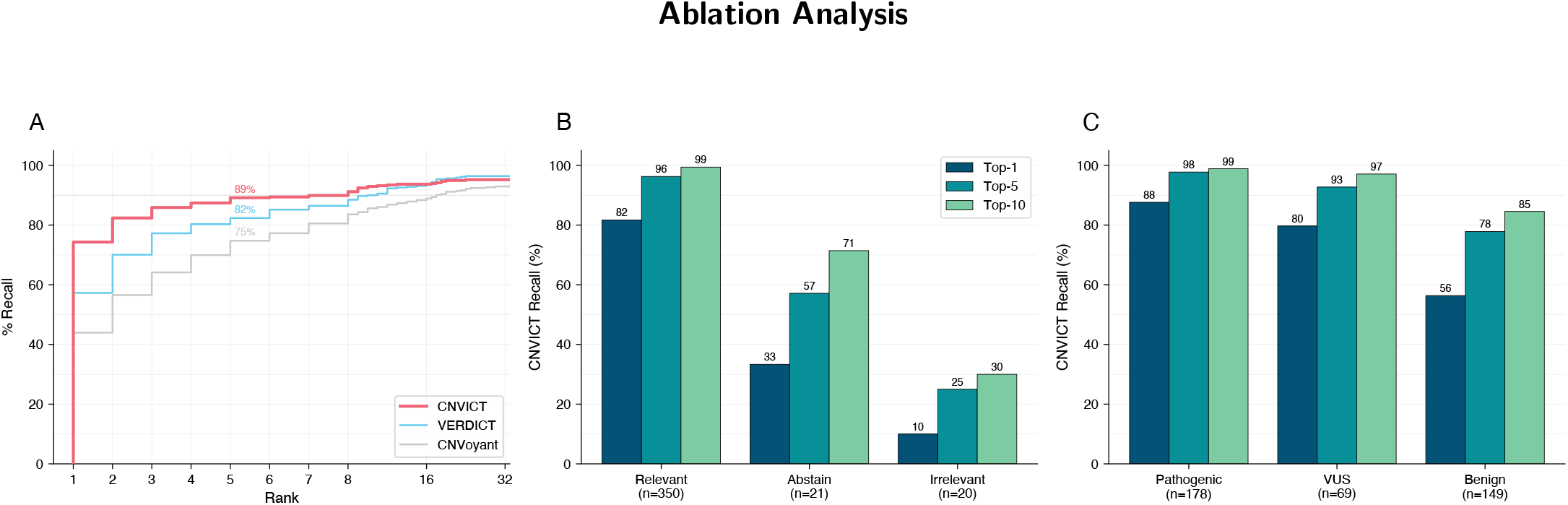
Ablation analysis and classification-stratified `CoNVict` performance. **(A)** Cumulative distribution of diagnostic CNV rank for `CoNVict` versus its two component modules: CNVerdict (phenotype relevance classifier) and CNVoyant (pathogenicity predictor). Annotations at rank 5 indicate the percentage of patients whose diagnostic CNV was ranked within the Top-5 by each tool. An inline legend identifies the three ablation curves. **(B)** `CoNVict` recall at Top-1, Top-5, and Top-10 stratified by CNVerdict classification (Relevant, Abstain, Irrelevant). Grouped bars show the percentage of diagnostic CNVs ranked at or above each threshold. Numeric values are annotated above each bar. Sample sizes per category are shown on the *x*-axis. **(C)** `CoNVict` recall at Top-1, Top-5, and Top-10 stratified by CNVoyant prediction (Pathogenic, VUS, Benign). Layout mirrors panel B. This figure demonstrates that `CoNVict` maintains robust recall even when its component classifiers disagree with the diagnostic label, though performance is highest when both components agree. See Supplementary Table S12 for Top-1/5/10/16/32/64 recall for `CoNVict`, CNVerdict, and CNVoyant.

**Table S13.**
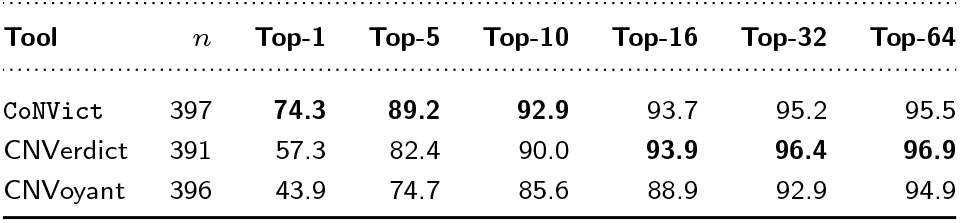
Top-*K* recall for ablation analysis. Top-1, Top-5, Top-10, Top-16, Top-32, and Top-64 recall (%) for `CoNVict` and its two component modules: CNVerdict and CNVoyant (pathogenicity predictor). CNVerdict was evaluated on *n* = 391 patients (6 patients excluded due to missing phenotype data); CNVoyant on *n* = 396 (1 patient excluded). Accompanies Supplementary Figure S11.

### Comparison with Phenotype-Agnostic Methods

### Ablation Analysis

### Simulation Benchmark Methodology

This section provides an expanded description of the simulation benchmark construction summarised in the main text. The benchmark comprises 397 simulated diagnostic cases, each containing a single causal copy-number variant (CNV) spiked into a realistic background of population-level structural variants. Cases were split into 320 whole-exome sequencing (WES) and 77 whole-genome sequencing (WGS) simulations across six CNV source categories.

#### Data Sources

The simulation pipeline draws on the following public resources:

- **ClinVar** (Landrum et al., 2014) (release 2026-02-08) — Pathogenic, Likely Pathogenic, and VUS copy-number variant entries exported as tab-separated files.
- **HPO Annotations** (Köhler et al., 2021) (version 2026-01-08) — Maps 8,576 OMIM diseases to Human Phenotype Ontology terms with frequency data.
- **1000 Genomes Phase 3 SV calls** (Sudmant et al., 2015; The 1000 Genomes Project Consortium, 2015) **—** Population-level structural variant calls (v8) from 2,504 samples, lifted over from GRCh37 to GRCh38 coordinates using the UCSC liftOver chain file.
- **DECIPHER CNV Syndromes** (Bragin et al., 2014) — 65 curated genomic disorder regions with GRCh38 coordinates, OMIM disease associations, and key genes (accessed February 2026).
- **GENCODE v44** (Frankish et al., 2021) — Gene and exon coordinates for approximately 20,000 protein-coding genes (GRCh38).
- **ClinGen Dosage Sensitivity** (Rehm et al., 2015) (accessed February 2026) — Haploinsufficiency (HI) and triplosensitivity (TS) scores for 1,624 genes and 505 curated genomic regions.
- **Undiagnosed Diseases Network (UDN)** (Ramoni et al., 2017; Alsentzer et al., 2023) — Validated gene–disease associations with clinician-curated phenotype terms and distractor gene lists.

In addition to the simulation data sources above, the annotation stage relies on AnnotSV (Geoffroy et al., 2018, 2021), which integrates population allele frequencies from gnomAD-SV (Collins et al., 2020), the Database of Genomic Variants (DGV) (MacDonald et al., 2014), and dbVar (Lappalainen et al., 2013), alongside pathogenicity and disease annotations from ClinVar, OMIM (Amberger et al., 2019), and ClinGen.

#### CNV Source Categories

Causal CNVs were drawn from six source categories:

1. **ClinVar Pathogenic (***n* = 60). Entries classified as Pathogenic, each ≥500 bp, with gene annotation and a mapped OMIM disease possessing HPO terms. A global cap of 30 entries each was enforced for 2-star and 3-star review status entries across all ClinVar sources to prevent over-representation of highly reviewed variants.
2. **ClinVar Likely Pathogenic (***n* = 60). Same filtering criteria and star-rating caps as Pathogenic.
3. **ClinVar VUS (***n* = 55). Variants of Uncertain Significance passing the same size, gene, and OMIM filters. The lower target reflects the reduced pass rate of VUS entries lacking disease associations.
4. **DECIPHER Syndromes (***n* = 55). Curated CNV syndrome regions representing well-characterised genomic disorders (e.g. DiGeorge syndrome, Williams–Beuren syndrome, Wolf–Hirschhorn syndrome). Each entry provides GRCh38 coordinates, an associated OMIM disease, a key gene, and an SV type (DEL or DUP).
5. **UDN (***n* = 60). Thirty cases used genes validated by the UDN that are absent from both ClinVar and ClinGen (novel gene–disease associations); the remaining 30 used UDN-validated genes with prior database annotations (10 each from ClinVar Pathogenic, Likely Pathogenic, and VUS tiers, with ClinGen-only genes filling shortfalls). CNVs were placed over each patient’s validated gene using size-category-based fabrication (Section 13.3). Each UDN patient additionally received distractor gene CNVs added to the background as realistic diagnostic noise.
6. **Computationally derived novel (***n* = 107). Seventy coding variants placed over ClinGen dosage-sensitive genes spanning all evidence tiers, restricted to genes with no existing structural variant ≥1 kb in ClinVar. The remaining 37 were non-coding variants in intronic or upstream regulatory regions verified against GENCODE v44 annotations (target was 40; three cases were dropped when placement could not satisfy the zero-exon-overlap constraint). All non-coding patients were assigned to WGS.

No variant identifier was reused across sources, and ClinVar entries underwent dual-pass loading: a first pass across all three significance files built a gene-to-OMIM mapping used as a fallback for entries lacking direct disease annotation.

#### Novel CNV Generation

##### Coding variants (*n* = 70)

The novel coding pool comprised ClinGen genes with haploinsufficiency or triplosensitivity score ≥1, excluding genes already present in ClinVar with a structural CNV ≥1 kb. Genes in the ClinVar structural set that were TS-eligible were retained as duplication-only targets. Duplications were capped at 50% of the target (35 maximum); the remaining slots were filled by deletions distributed across five size categories to ensure broad coverage of the clinical size spectrum:

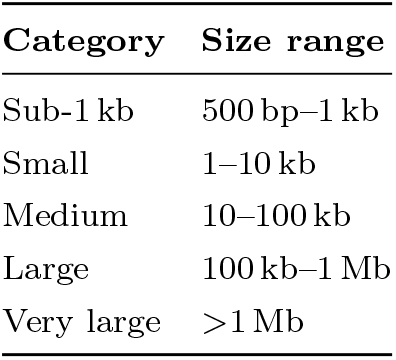

For each variant, the CNV was placed over the target gene according to the assigned size category: sub-kilobase variants were anchored to a random exon, small variants spanned one to three consecutive exons, medium variants extended 500–5,000 bp beyond gene boundaries, large variants were split asymmetrically around the gene, and very large variants were centred on the gene midpoint. After placement, Gaussian breakpoint noise (mean = 0, s.d. = 500 bp) was added independently to both breakpoints, with a minimum size guarantee of 500 bp. SV type (DEL or DUP) was determined by ClinGen dosage evidence: HI-eligible genes yielded deletions, TS-eligible genes yielded duplications.

##### Non-coding variants (*n* = 37)

Non-coding CNVs were placed near target genes without overlapping any protein-coding exon, using two strategies selected at random:

- **Intronic placement (60% probability):** The CNV was placed within the largest intron (≥2 kb) of the target gene, sized between 1 and 10 kb, with a 50 bp minimum buffer from exon boundaries.
- **Upstream regulatory placement (40% probability):** The CNV was placed 1–50 kb upstream of the transcription start site in a strand-aware manner (upstream of minus-strand genes corresponds to positions after the gene end), sized between 1 and 20 kb.

Both strategies verified zero exon overlap via binary search on merged coding exon intervals. Three patients were dropped when neither strategy could satisfy this constraint.

#### UDN patient CNVs

UDN patients used the same coding fabrication procedure, with the validated UDN gene as the target and size category determined from gene length. Distractor gene CNVs (one per distractor gene in the UDN record) were generated with random mechanism assignment (DEL or DUP with equal probability) and added to the background.

#### Breakpoint Perturbation

ClinVar and DECIPHER diagnostic CNVs underwent randomised breakpoint perturbation to prevent trivial identification via exact coordinate matching against source databases. One of four strategies was assigned at random according to configured weights:

1. **Exon shift (25%):** One breakpoint was shifted to exclude one to two exons from one end while including extra exons from the other, altering the exon count and biological severity assessment. This strategy required the target gene to have ≥3 exons with ≥2 overlapped by the original CNV.
2. **Neighbour extend (25%):** The closer breakpoint was extended to overlap at least one exon of the nearest neighbouring gene (within 500 kb), converting a single-gene CNV into a multi-gene event that tests multi-gene handling.
3. **Shrink to partial (20%):** The CNV was shrunk to cover only one to two exons (if ≥3 were originally overlapped) or to 30–60% of gene body coverage (if original coverage was ≥70% with ≥2 exons), reducing exon count and potentially changing whole-gene to intragenic classification.
4. **Classic (30%):** Each breakpoint was independently shifted by ±5–10% of the original CNV length, clamped to [200 bp, 5 kb]. Direction was randomised independently per breakpoint.

Each strategy included a fallback to the classic method when applicability conditions were not met. A noncoding guard check was applied after perturbation: if the original CNV was classified as non-coding (intronic or regulatory) and the perturbed coordinates overlapped a coding exon, the perturbation was reverted to the original coordinates.

#### Background Structural Variant Construction

Background CNV profiles were drawn from the 1000 Genomes Project Phase 3 structural variant calls, lifted over to GRCh38 coordinates and filtered to retain only PASS-flagged, precisely breakpointed variants (no IMPRECISE flag). Allele classification followed copy-number ALT encoding: CN0 alleles were classified as deletions, CN≥2 as duplications, and hemizygous-loss-only (CN1) alleles were excluded. QUAL-based filtering was intentionally omitted because all duplication variants in the 1000G call set carry a missing quality score (QUAL=‘.’), which would have eliminated all duplications from the background.

Each patient was assigned a unique 1000G sample (no reuse across the 397 patients). The 80 samples with the fewest total structural variants were reserved for WGS patients (mean 969 SVs per patient); the remaining ∼2,424 samples, sorted by proximity to 200 exon-overlapping SVs, were assigned to WES patients (mean 129 SVs per patient). In WES mode, only SVs overlapping protein-coding exons (unpadded intervals) were retained. Any background SV overlapping the diagnostic CNV or UDN distractor CNVs was removed to prevent confounding.

#### Clinical Narrative and Phenotype Generation

For each non-UDN patient, a synthetic clinical narrative was generated in three steps. First, the causal gene–disease pair was queried against Perplexity Sonar, an LLM-grounded search engine, to retrieve characteristic clinical features of the associated disorder, including common and rare symptoms, typical age of onset, inheritance pattern, and progression trajectory. Second, Gemini 2.5 Flash synthesized these retrieved features into a concise patient epicrisis resembling a realistic clinical vignette. Third, an HPO-based retrieval-augmented generation (RAG) system processed the epicrisis to extract up to five standardized Human Phenotype Ontology terms (Köhler et al., 2021), mapping free-text clinical descriptions to structured phenotype annotations. This pipeline captures clinical heterogeneity beyond database-curated HPO annotations, which often omit rare or atypical presentations.

UDN patients instead retained their original clinician-validated phenotype terms, with up to five terms randomly sampled from the patient’s positive phenotypes field.

#### Development Set

A separate subset of 38 cases, constructed using the same methodology and source distribution, was reserved for pipeline development and prompt tuning. These cases were excluded from all reported evaluations.

### Gene Cache Prompts & Analysis

**Table S14.** Overview of the gene knowledge cache used for keyword-based phenotype matching.

| Property | Value |
| --- | --- |
| Total genes in cache | 47,852 |
| Genes with ≥ 1 keyword | 31,099 (65.0%) |
| Genes with 0 keywords | 16,753 (35.0%) |
| Total keywords | 455,122 |

**Table S15.**
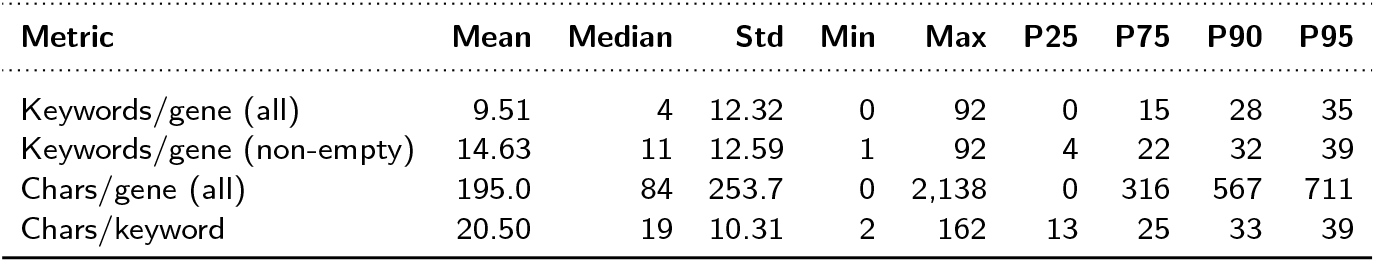
Distribution statistics of the gene knowledge cache. “Keywords/gene (non-empty)” excludes the 16,753 genes with no associated keywords.

| Metric | Mean | Median | Std | Min | Max | P25 | P75 | P90 | P95 |
| --- | --- | --- | --- | --- | --- | --- | --- | --- | --- |
| Keywords/gene (all) | 9.51 | 4 | 12.32 | 0 | 92 | 0 | 15 | 28 | 35 |
| Keywords/gene (non-empty) | 14.63 | 11 | 12.59 | 1 | 92 | 4 | 22 | 32 | 39 |
| Chars/gene (all) | 195.0 | 84 | 253.7 | 0 | 2,138 | 0 | 316 | 567 | 711 |
| Chars/keyword | 20.50 | 19 | 10.31 | 2 | 162 | 13 | 25 | 33 | 39 |

**Table S16.** Distribution of keyword counts per gene in the knowledge cache.

| Keywords per gene | Genes | % |
| --- | --- | --- |
| 0 | 16,753 | 35.0 |
| 1–5 | 9,214 | 19.3 |
| 6–10 | 5,694 | 11.9 |
| 11–20 | 7,620 | 15.9 |
| 21–50 | 8,149 | 17.0 |
| 51–100 | 422 | 0.9 |

**Figure S17.**
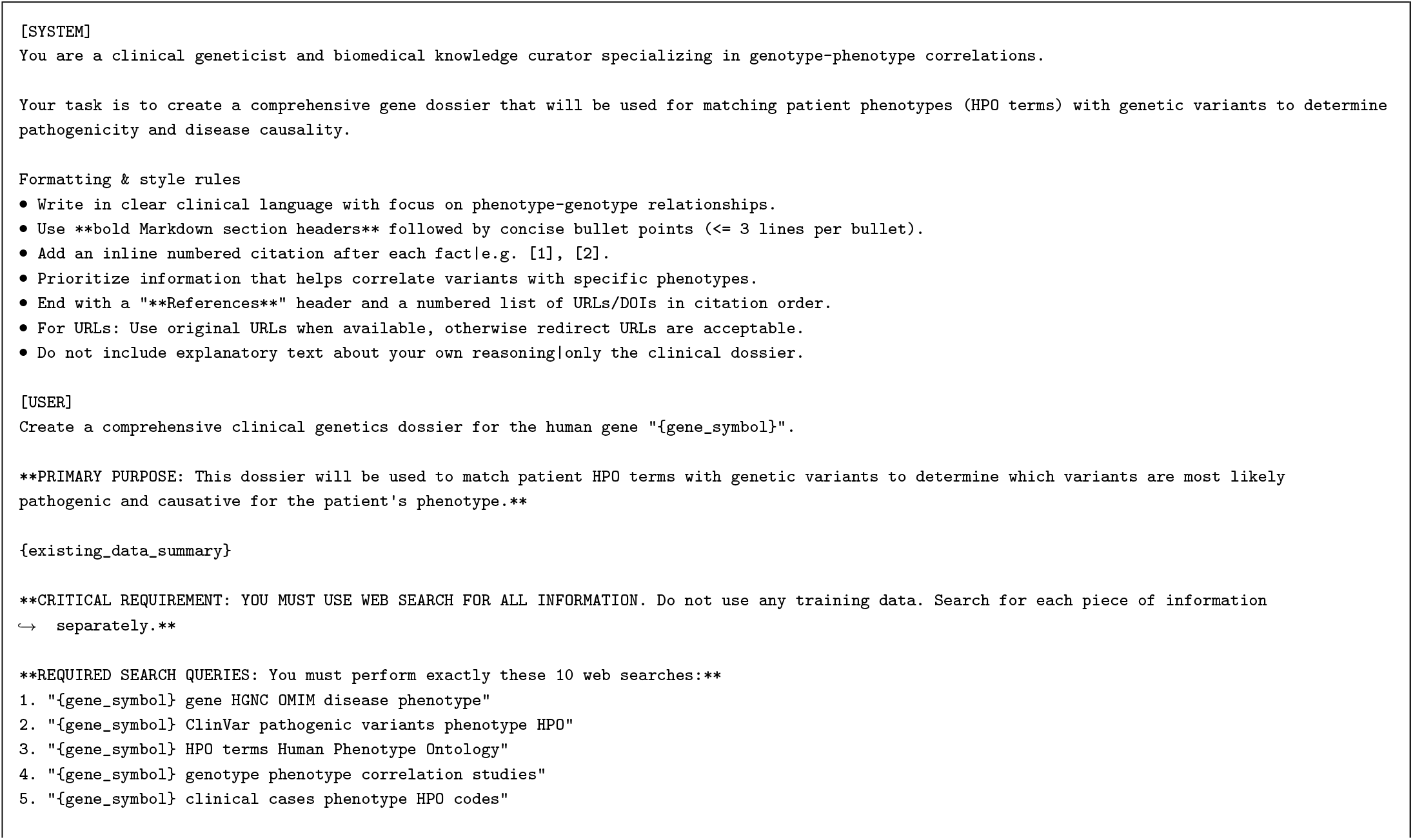
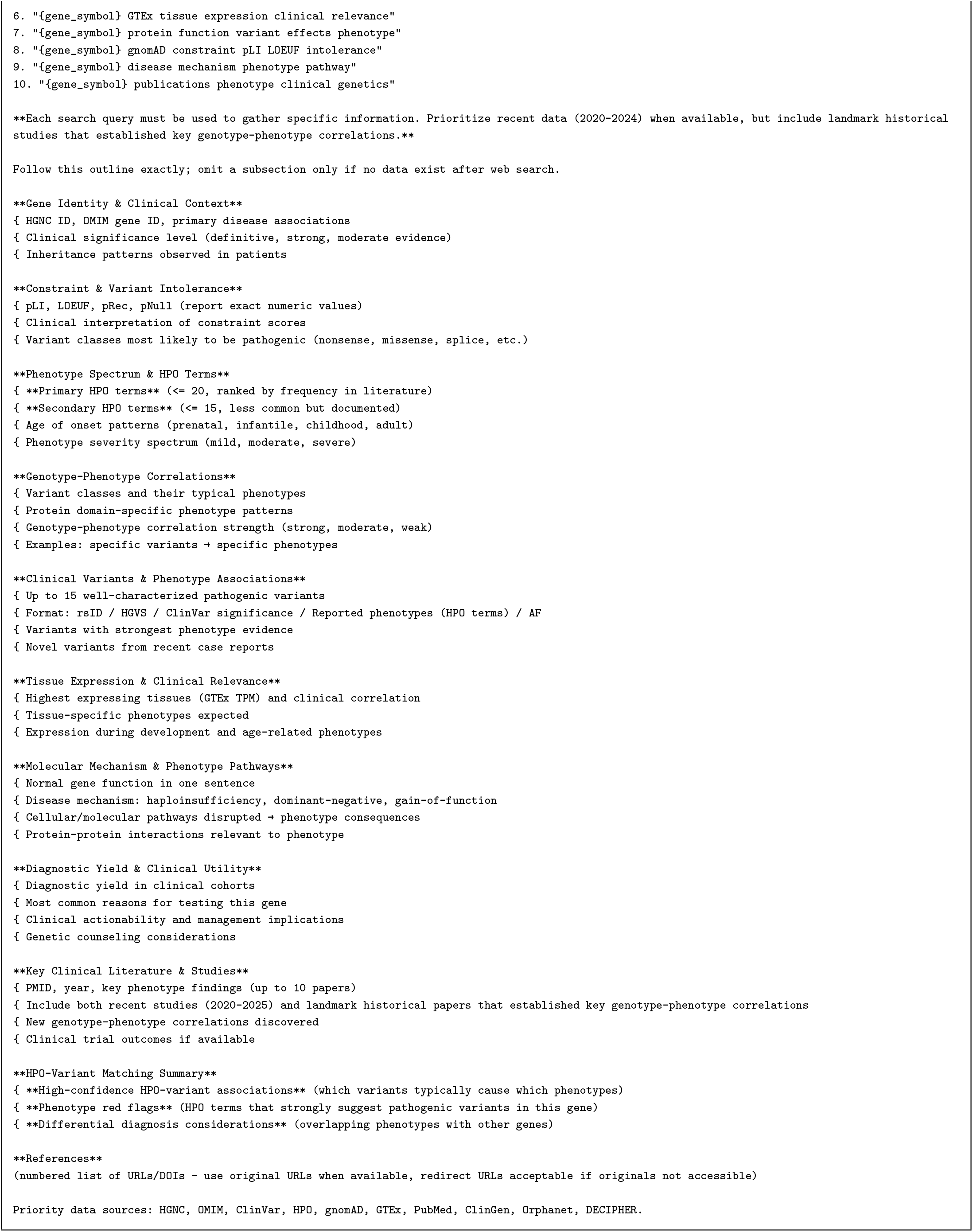
Prompt template for LLM-based gene dossier generation with grounded web search. (gene symbol) is replaced per gene and (existing data summary) is populated with structured dbNSFP fields when available. The prompt is sent to Gemini 2.0 Flash with the Google Search tool enabled.

**Figure S18.**
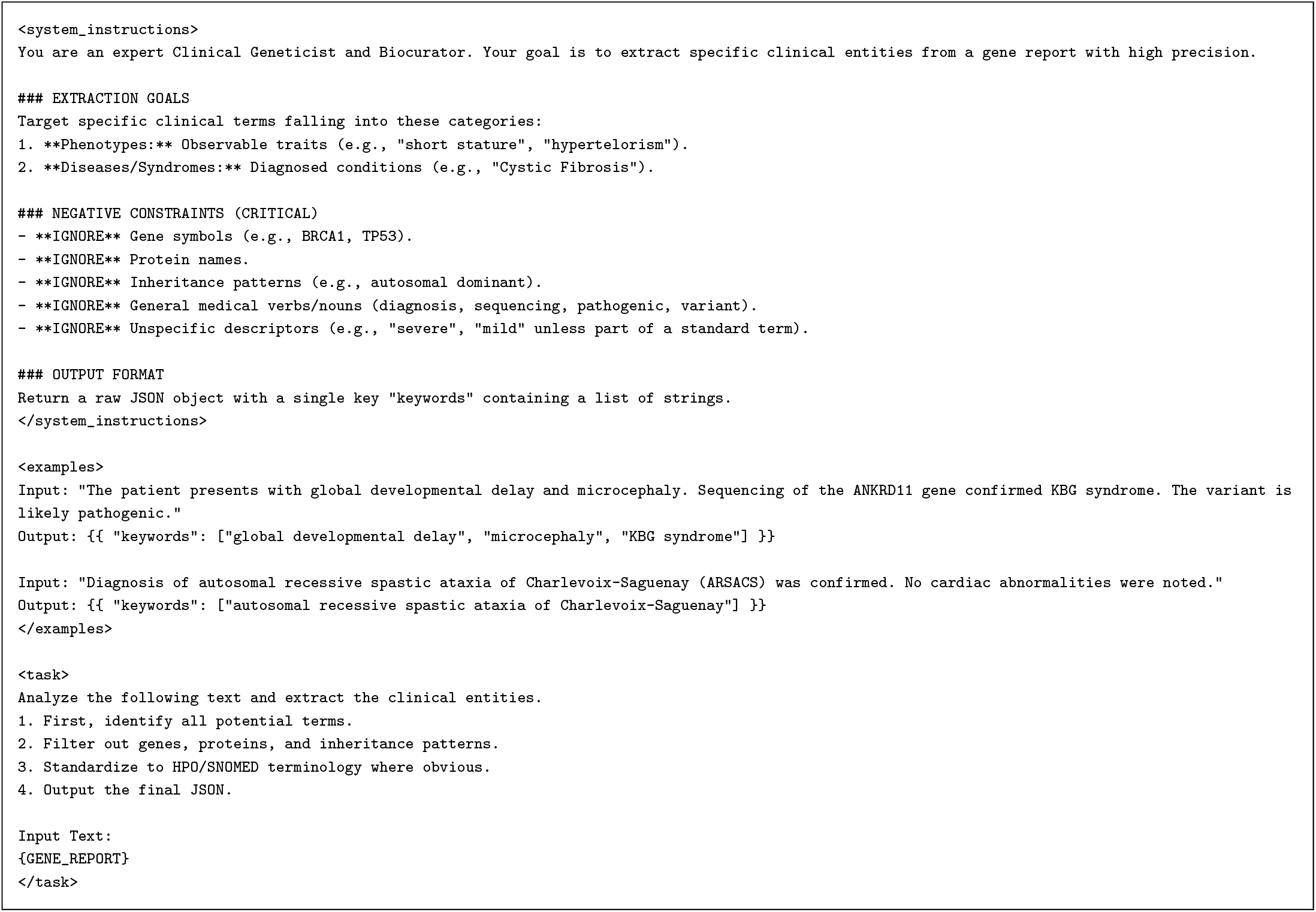
Prompt template for LLM-based gene keyword extraction from gene dossiers. (GENE REPORT) is replaced with gene dossiers. The prompt is sent to Gemini 2.5 Flash Lite.

### Filtering Approaches

**Table S17.** Dataset overview. Gene-bearing CNVs are those overlapping at least one protein-coding or regulatory gene.

| Metric | WES | WGS | All |
| --- | --- | --- | --- |
| Samples | 320 | 77 | 397 |
| Total CNVs | 42,262 | 74,739 | 117,001 |
| CNVs/sample (median) | 131 | 974 | 134 |
| CNVs/sample (mean) | 132.1 | 970.6 | 294.7 |
| Gene-bearing CNVs | 41,333 | 39,120 | 80,453 |
| No-gene CNVs | 929 | 35,619 | 36,548 |
| % gene-bearing | 97.8 | 52.3 | 68.8 |
| Gene-bearing/sample (mean) | 129.2 | 508.1 | 202.7 |
| Genes/CNV mean (gene-bearing) | 1.8 | 1.3 | 1.6 |

**Table S18.** Pre-filter ablation: target CNV recall (%) at selected top-*k* cutoffs for CNVoyant and AnnotSV Score ranking. Approximate mean LLM calls per sample (CNVerdict + Tournament) shown in parentheses. “No filter” passes all gene-bearing CNVs. Accompanies Supplementary Figure S19.

| Top-*k* | WES (*n*=320) |  | WGS (*n*=77) |  |
| --- | --- | --- | --- | --- |
|  | CNVoyant | AnnotSV | CNVoyant | AnnotSV |
| 8 | 89.7 (~21) | 82.8 (~21) | 57.1 (~8) | 70.1 (~8) |
| 16 | 94.4 (~41) | 90.9 (~41) | 64.9 (~53) | 74.0 (~56) |
| 32 | 98.1 (~78) | 98.1 (~78) | 70.1 (~100) | 74.0 (~105) |
| 64 | 99.1 (~110) | 99.7 (~110) | 76.6 (~171) | 89.6 (~180) |
| 128 | 99.7 (~169) | 100.0 (~170) | 84.4 (~229) | 100.0 (~234) |
| 512 | 100.0 (~175) | 100.0 (~175) | 96.1 (~443) | 100.0 (~599) |
| No filter | 100.0 (~175) |  | 100.0 (~629) |  |

**Figure S19.**
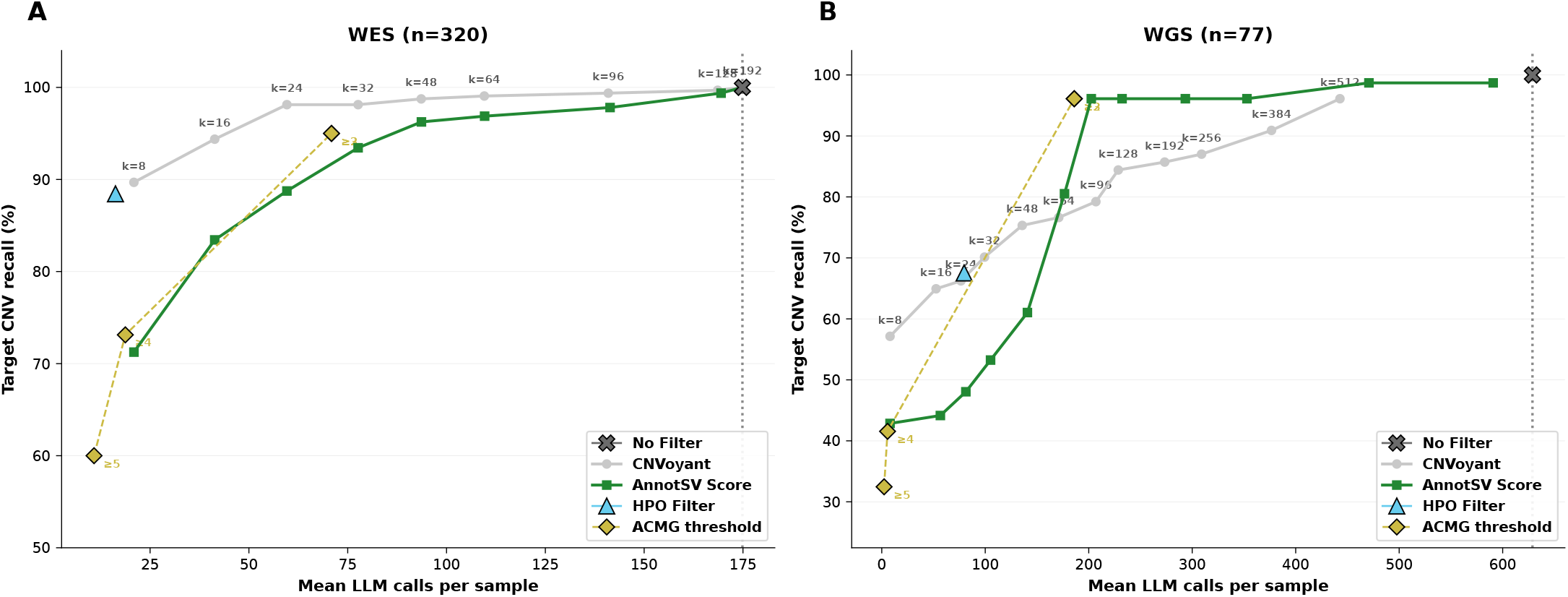
Pre-filter cost–recall trade-off. Each point represents a top-*k* cutoff (labelled), with the *x*-axis showing approximate mean LLM call count per sample (CNVerdict + Tournament) and the *y*-axis showing target CNV recall. The dashed vertical line marks the no-filter baseline. CNVoyant and AnnotSV Score ranking strategies are compared alongside HPO annotated gene filtering and ACMG class (1-5) thresholds. **(A)** WES, **(B)** WGS.

### CNVerdict and Tournament

#### CNVerdict and Tournament Analyses

**Table S19.**
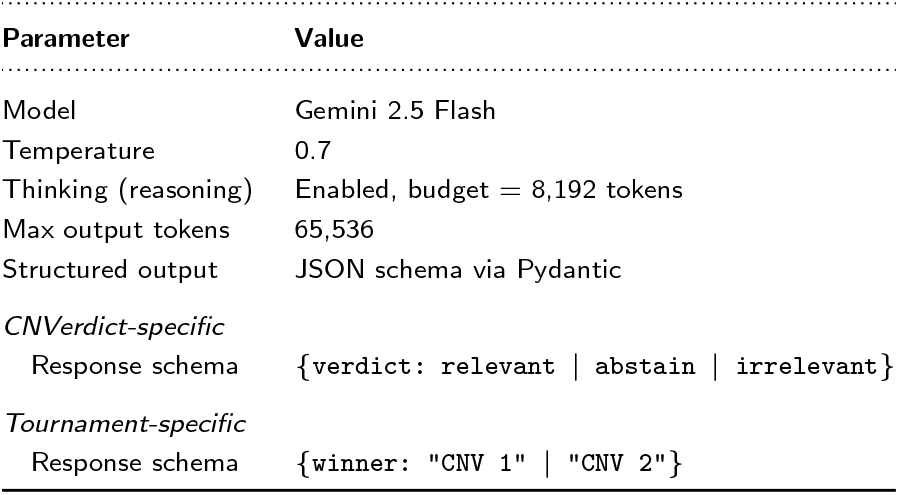
LLM configuration used for both the CNVerdict and Tournament stages. Both stages share the same model and generation parameters; they differ only in prompt template and structured output schema.

**Table S20.** LLM call counts across the full benchmark (*n*_WES_ = 320, *n*_WGS_ = 77, *N* = 397 samples). CNVerdict calls equal the pre-filtered CNV count per sample (64 for WES, 512 for WGS); CNVs with zero genes are auto-classified without an LLM call and are excluded from the totals below. Tournament calls are pairwise comparisons within the single-elimination-plus-extraction algorithm.

| Stage | Cohort | Input | Output | Calls/sample | Samples | Total calls |
| --- | --- | --- | --- | --- | --- | --- |
| CNVerdict | WES | 64 | — | 64 | 320 | 20,480 |
|  | WGS | 512 | — | 512 | 77 | 39,424 |
| Tournament | WES | top-32 | top-4 | 48.5 (mean) | 320 | 15,516 |
|  | WGS | top-64 | top-8 | 143.5 (mean) | 77 | 11,047 |
| Grand total |  |  |  |  | 397 | 86,467 |

**Table S21.**
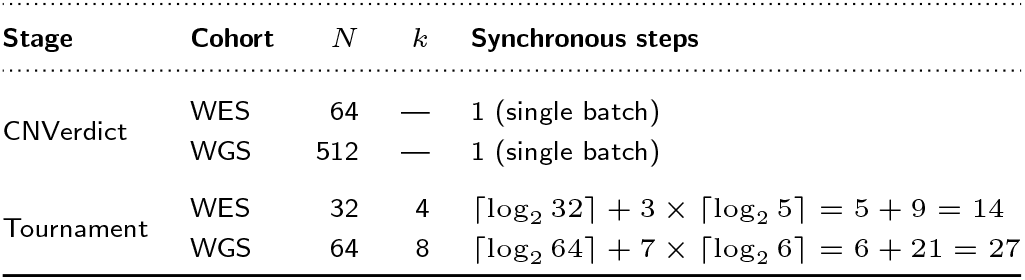
Parallel (synchronous) step analysis per sample. Within each step, all LLM calls execute concurrently. The CNVerdict stage issues all classification requests in a single batch. The Tournament stage first builds a single-elimination tree in ⌈log_2_⌉ *N* rounds, then sequentially extracts ranks 2 through *k* via sub-tournaments over the pool of candidates that lost to already-selected winners.

| Stage | Cohort | $N$ | $k$ | Synchronous steps |
| --- | --- | --- | --- | --- |
| CNVerdict | WES | 64 | — | 1 (single batch) |
|  | WGS | 512 | — | 1 (single batch) |
| Tournament | WES | 32 | 4 | $\lceil \log_2 32 \rceil + 3 \times \lceil \log_2 5 \rceil = 5 + 9 = 14$ |
| | WGS | 64 | 8 | $\lceil \log_2 64 \rceil + 7 \times \lceil \log_2 6 \rceil = 6 + 21 = 27$ |

#### Clinical Context

Each CNV is serialised into a Markdown-formatted text block before being inserted into the prompt template. The representation consists of two parts:

1. **CNV header**. Genomic coordinates, variant type, size and size category, cytoband, gene count, and full-row AnnotSV annotations (ACMG class, AnnotSV ranking score, pathogenic-gain/loss phenotypes and HPO matches, population allele frequency, and caller quality metrics when available).
2. **Per-gene blocks**. For every gene overlapping the CNV, the block includes up to 15 phenotype keywords from the gene knowledge cache (Table S14) and split-level AnnotSV annotations: pLI, LOEUF bin, ClinGen haploinsufficiency/triplosensitivity class, DDD HI percentile, OMIM phenotype and inheritance, GenCC disease and mode of inheritance, transcript location, overlapped CDS length and percentage, and ExAC deletion/duplication *z*-scores. In the CNVerdict stage, *all* overlapping genes are listed. In the Tournament stage, only genes that have at least one keyword in the knowledge cache are shown, and an additional **Regulatory genes** section lists genes whose enhancers or regulatory elements overlap the CNV (annotated by AnnotSV’s RE gene field).

The **patient context** block, shared by both stages, contains the patient’s epicrisis (free-text clinical summary) and a list of HPO terms enriched with their definitions.

#### CNVerdict and Tournament Contexts

**Figure S20.**
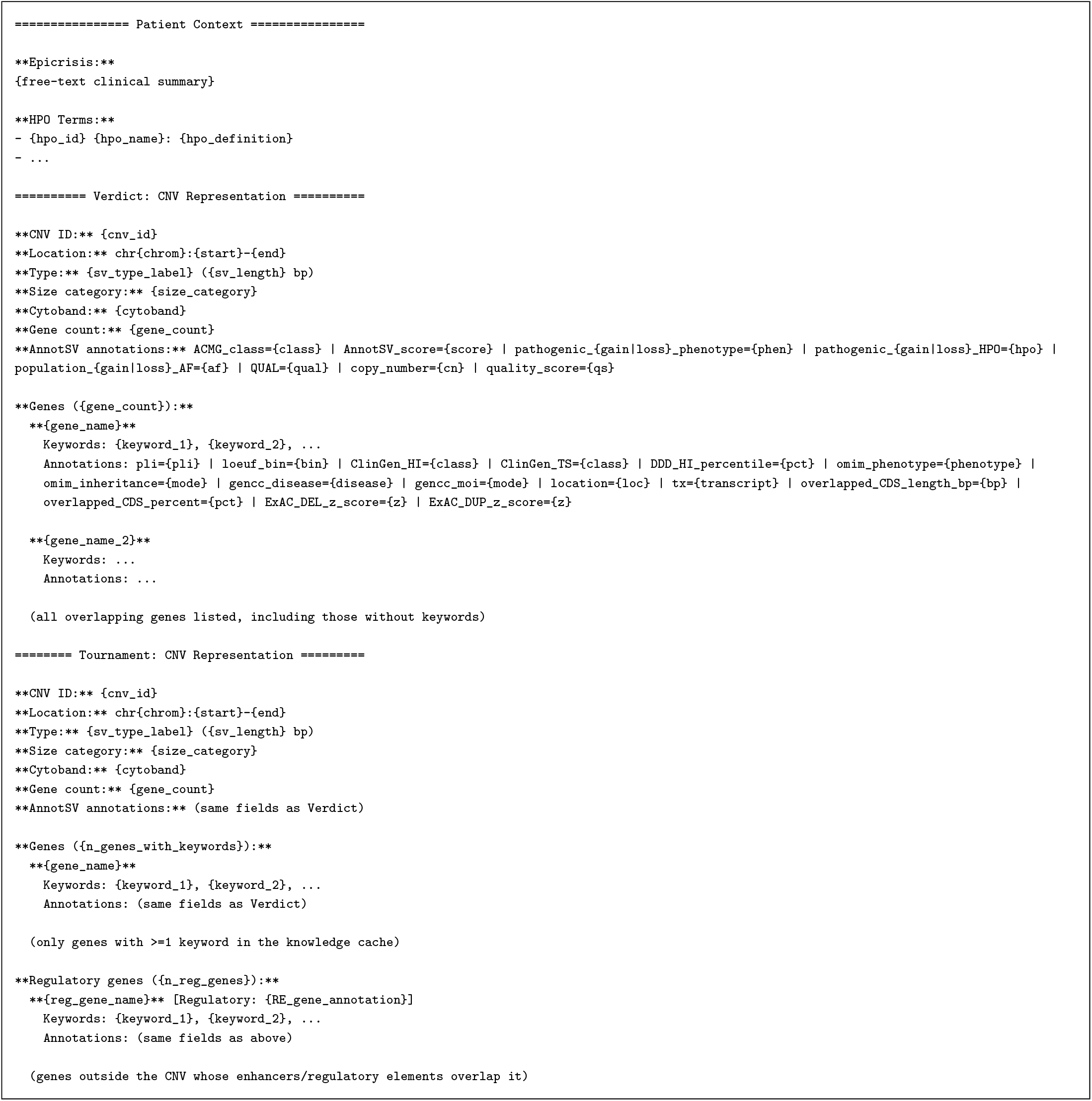
Template of the serialised patient context and CNV representation passed to the LLM. The top section shows the patient context format (epicrisis and enriched HPO terms with definitions). The middle section shows the Verdict format: all overlapping genes are listed with phenotype keywords and AnnotSV gene-level annotations. The bottom section shows the Tournament format: only genes with at least one keyword in the knowledge cache are included, and a separate **Regulatory genes** section lists genes whose enhancers or regulatory elements overlap the CNV. Annotation fields are included only when non-empty.

**Figure S21.**
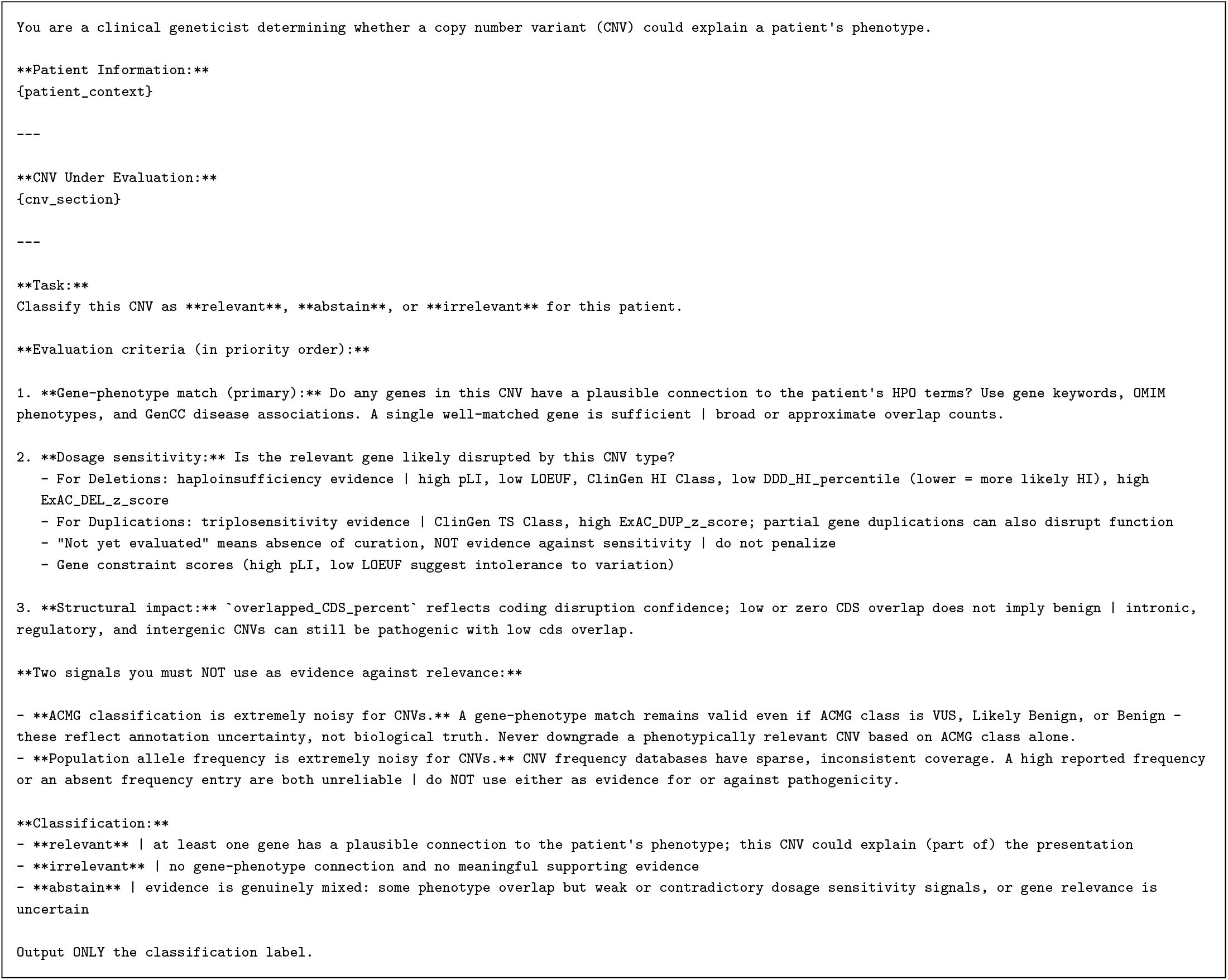
CNVerdict classification prompt template. `{patient_context}` is replaced with the patient’s epicrisis and enriched HPO terms; `{cnv_section}` contains the CNV header (location, type, size, cytoband, gene count, AnnotSV annotations) and per-gene blocks with phenotype keywords and gene-level annotations. The LLM returns a structured response constrained to `relevant, abstain`, or `irrelevant`.

**Figure S22.**
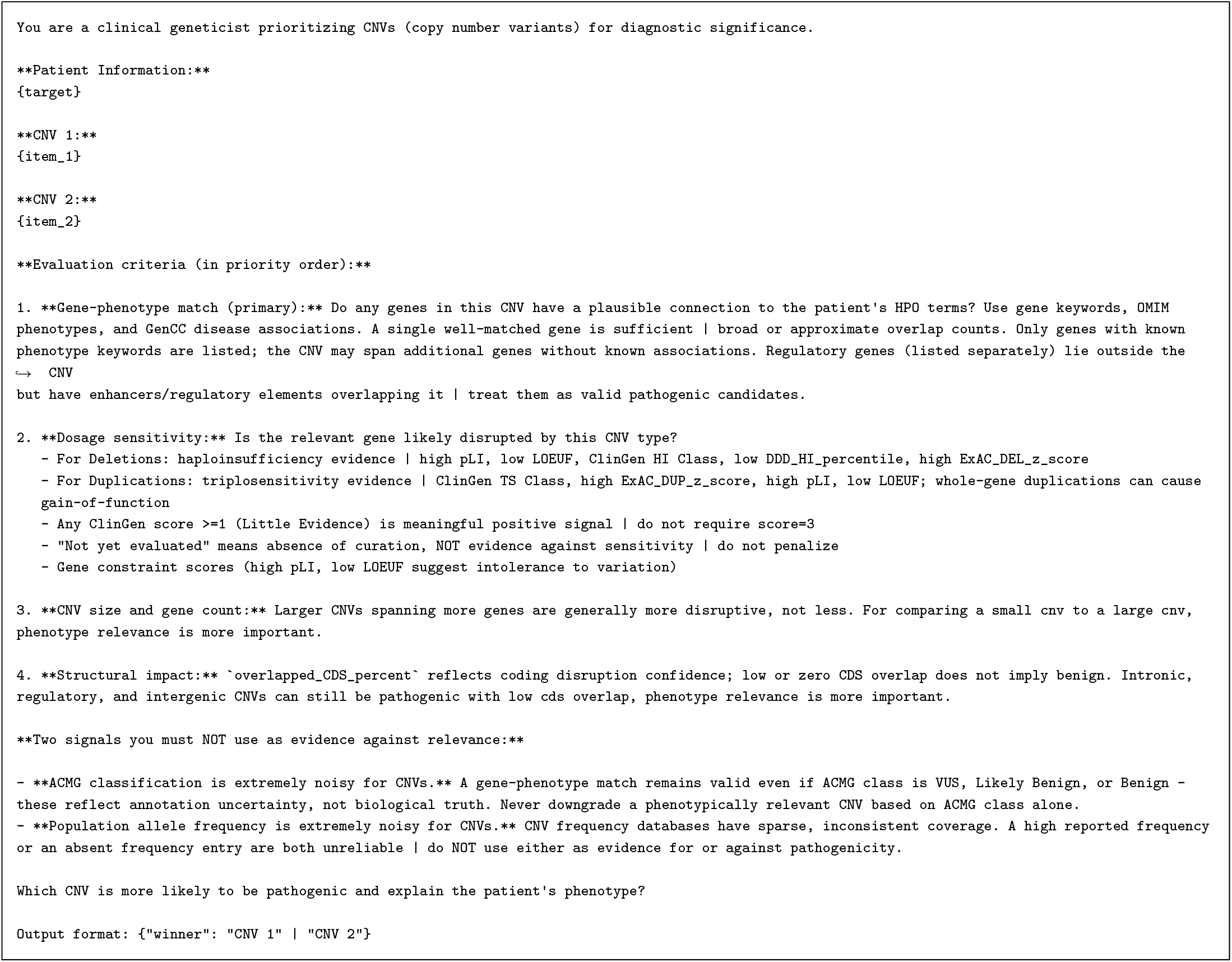
Tournament pairwise comparison prompt template. `{target}` is replaced with the patient context; `{item_1}` and `{item_2}` are two candidate CNVs formatted with gene keywords, AnnotSV annotations, and regulatory gene information. The LLM returns a structured JSON response indicating the winner of the pairwise comparison.

